# Host immunological responses facilitate development of SARS-CoV-2 mutations in patients receiving monoclonal antibody treatments

**DOI:** 10.1101/2022.09.20.22280135

**Authors:** Akshita Gupta, Angelina Konnova, Mathias Smet, Matilda Berkell, Alessia Savoldi, Matteo Morra, Vincent Van averbeke, Fien De Winter, Denise Peserico, Elisa Danese, An Hotterbeekx, Elda Righi, mAb ORCHESTRA working group, Pasquale De Nardo, Evelina Tacconelli, Surbhi Malhotra Kumar, Samir Kumar Singh

## Abstract

The role of host immunity in emergence of evasive SARS-CoV-2 Spike mutations under therapeutic monoclonal antibody (mAb) pressure remains to be explored. Here, we show that patients treated with various anti-SARS-CoV-2 mAb regimens develop evasive Spike mutations with remarkable speed and high specificity to the targeted mAb-binding sites. Mutations develop more frequently in immunocompromised patients and strongly correlate not only with the neutralizing capacity of the therapeutic mAbs, but also with an anti-inflammatory and healing-promoting host milieu. Machine-learning models based on soluble host-derived biomarkers identified patients at high risk of developing escape mutations against therapeutic mAbs with high accuracy. While our data demonstrate that host-driven immune and non-immune responses are essential for development of mutant SARS-CoV-2, these data could also support point-of-care decision making in reducing the risk of mAb treatment failure and improving mitigation strategies for possible dissemination of escape SARS-CoV-2 mutants.

## Introduction

The coronavirus replication machinery encodes proofreading functions resulting in fewer errors compared to other RNA viruses, however, multiple SARS-CoV-2 variants of concern (VOCs) have emerged throughout the pandemic carrying VOC-defining mutations. For example, Alpha (B.1.1.7), Beta (B.1.351), Gamma (P.1), Delta, Zeta, Eta, Theta, Iota, and Omicron (BA.1/BA.2) variants have been shown to carry a distinct set of mutations, which evade existing natural neutralizing antibody responses (1-3)(reviewed in ref (4)).

SARS-CoV-2 mutation rates are higher in immunocompromised or severely ill patients who show prolonged SARS-CoV-2 infections or carriage (5-12). Immunocompromised individuals are also unable to develop sufficient antibody titers after the administration of SARS-CoV-2 vaccines. To tackle this, synthetic anti-SARS-CoV-2 neutralizing monoclonal antibodies (mAbs) targeting the Spike protein have been developed that demonstrate clinical benefit for mild-to-moderately ill, high-risk COVID-19 patients (13-20). For example, the first widely available mAb, bamlanivimab that targets an epitope on the receptor-binding domain (RBD), showed a reduced rate of hospitalization, ICU admission, and mortality compared with usual care (21). The addition of etesevimab to bamlanivimab resulted in improved clinical outcomes due to overlapping binding epitopes within Spike RBD, concomitant to the emergence of SARS-CoV-2 VOCs, mainly B.1.351 and P.1 (22). The success of combination mAb therapy led to use of casirivimab/imdevimab, with distinct binding sites in the Spike RBD region, in at-risk populations, resulting in decreased rate of hospitalization (23). As the pandemic evolved and new VOCs were identified, sotrovimab was developed with a modified Fc domain along with an increased half-life (13, 14). These modifications target highly conserved epitopes on Spike causing conformational transitions necessary for association with the ACE2 receptor (15), resulting in reduced risk of disease progression and death (13, 15).

Several reports have also identified *de novo* mutations under therapeutic mAb pressure, including E484Q/K and Q493K/R under bamlanivimab/etesevimab pressure (24-26) and P337R/S, E340D/K/V, and G446S/V under casirivimab/imdevimab and/or sotrovimab pressure (27-30). However, despite the widespread use of mAbs, these studies are rather few, and were conducted in limited number of patients. Moreover, the role of host immune pressure in selection of mAb-driven *de novo* SARS-CoV-2 mutations has not been explored so far.

Here, we characterize the development of SARS-CoV-2 mutations in patients treated with bamlanivimab, bamlanivimab/etesevimab, casirivimab/imdevimab, or sotrovimab in relation to their neutralization potential against SARS-CoV-2 VOCs. We focus on natural humoral and cellular host immunity including responses mediated by cytokines and other correlates of adaptive evolution.

## Results

### Immunocompromised COVID-19 patients receiving early mAb therapy continue to display significantly higher viral loads compared to non-immunocompromised patients

The H2020-funded ORCHESTRA project (Connecting European Cohorts to Increase Common and Effective Response to SARS-CoV-2 Pandemic) includes work package 2 (WP2), prospectively enrolling high-risk patients receiving early treatment for symptomatic COVID-19. Clinical efficacies of bamlanivimab, bamlanivimab/etesevimab, casirivimab/imdevimab, or sotrovimab in 740 mild-to-moderate non-hospitalized COVID-19 patients have been described (19, 20) (for eligibility criteria, see **Supplementary Table 1**). From the WP2 cohort, patients were prospectively invited to a sub-study assessing immunological and virological responses to mAbs. Overall, 204 patients were enrolled receiving bamlanivimab (n = 45), bamlanivimab/etesevimab (n = 108), casirivimab/imdevimab (n = 17), or sotrovimab (n = 34) (**Table 1**). Patients were assessed and sampled before mAb infusion (D0) and after treatment at day (D)2, D7, and in 98 patients at D28. The maximum study length of 28 days was chosen as the mean duration of SARS-CoV-2 RNA shedding from the upper respiratory tract has been estimated as not more than 17 days (31, 32). Patient groups did not differ significantly in WHO progressive severity score (33). The median age of the total study cohort was 64 years (inter-quartile range (IQR): 57-75) and 54.2% of the enrolled patients were males. During the 28-day follow-up, 28 patients (28/204; 13.7%) were hospitalized for severe COVID-19 (bamlanivimab: 8/45 (17.7%); bamlanivimab/etesevimab: 20/108 (18.5%)) and 3/204 patients died (1.5%). For patient characteristics, see Table 1.

**Table 1.** Patient characteristics of enrolled patients treated with bamlanivimab, bamlanivimab/etesevimab, casirivimab/imdevimab, or sotrovimab in the study. IQR: inter quartile range. mo: months.

| Patient characteristics | Bamlanivimab (N=45) | Bamlanivimab/etesevimab (N=108) | Casirivimab/imdevimab (N=17) | Sotrovimab (N=34) | P-value |
| --- | --- | --- | --- | --- | --- |
| Male (%) | 30 (66.7) | 59 (54.6) | 9 (52.9) | 13 (37.1) | NS |
| Age (median, IQR) | 63 (58-78) | 65 (58-75) | 53 (47-63) | 69 (61-76) | 0.03 |
| < 65 years | 58 (52-61) | 58 (51-62) | 52 (47-62) | 58 (50-63) | NS |
| ≥ 65 years | 78 (72-83) | 75 (70-78) | 76 (74-77) | 75 (70-79) | NS |
| BMI (median, IQR) | 28 (24-31) | 29 (25-35.25) | 29 (26-36) | 28 (25-30) | NS |
| Hospital admission (%) | 8 (17.8) | 20 (18.5) | 0 (0.0) | 0 (0.0) | 0.012 |
| Death (%) | 2 (4.4) | 1 (0.9) | 0 (0.0) | 0 (0.0) | NS |
| WHO progression severity scale – At enrolment (median, IQR) | 2 (2-2) | 2 (2-2) | 2 (2-2) | 3 (3-3) | NS |
| WHO progression severity scale – Worst (median, IQR) | 6 (4.75-7) | 4 (4-5.25) | 2 (2-2) | 3 (3-3) | <0.001 |
| Days from symptoms onset to mAb infusion (median, IQR) | 6 (4-7) | 5 (4-6) | 6 (5-7) | 3 (2-4) | <0.001 |
| sO<sub>2</sub> % (median, IQR) | 95 (94-97) | 97 (95-98) | 97 (97-98) | 97 (96-98) | <0.001 |
| Anti-SARS-CoV-2 vaccination (>2 weeks post-dose, ≥ 2 doses) | 0 (0.0) | 0 (0.0) | 1 (0.9) | 25 (73.5) | <0.001 |
| **Ongoing COVID-related therapy** |  |  |  |  |  |
| Prednisone (%) | 5 (11.1) | 19 (17.6) | 3 (17.6) | 0 (0.0) | <0.001 |
| Azithromycin (%) | 3 (6.7) | 10 (9.3) | 1 (5.9) | 0 (0.0) | NS |
| Amoxicillin/clavulanate (%) | 1 (2.2) | 6 (5.6) | 2 (11.8) | 0 (0.0) | NS |
| **Comorbidities** |  |  |  |  |  |
| Diabetes (with or without damage) (%) | 8 (17.8) | 20 (18.5) | 2 (11.8) | 5 (14.3) | NS |
| Cardiovascular disease (ischemic/arrhythmia/Hypertension) (%) | 12 (26.7) | 45 (41.7) | 6 (35.3) | 14 (40.0) | 0.012 |
| Hematologic cancer (with ongoing CHT/ongoing stopped < 6 mo) (%) | 0 (0.0) | 1 (0.9) | 0 (0.0) | 8 (22.8) | <0.001 |
| Solid organ cancer (with ongoing therapy/ongoing stopped < 6 mo) (%) | 0 (0.0) | 0 (0.0) | 0 (0.0) | 4 (11.4) | <0.001 |
| Chronic renal failure (with or without need of dialysis) (%) | 2 (4.4) | 5 (4.6) | 1 (5.9) | 4 (11.4) | NS |
| Chronic pulmonary diseases (%) | 10 (22.2) | 18 (16.7) | 4 (23.5) | 7 (20.0) | NS |
| Any neurological/vascular disease (%) | 3 (6.7) | 9 (8.3) | 0 (0.0) | 1 (2.9) | NS |
| Transplant recipients (%) | 0 (0.0) | 0 (0.0) | 0 (0.0) | 4 (11.4) | <0.001 |
| Immunological diseases requiring immunosuppressive agents (%) | 3 (6.7) | 11 (10.2) | 1 (5.9) | 15 (42.8) | <0.001 |
| **Symptoms** |  |  |  |  |  |
| Anosmia (%) | 3 (6.7) | 2 (1.9) | 3 (17.6) | 0 (0.0) | 0.007 |
| Ageusia (%) | 5 (11.1) | 7 (6.5) | 2 (11.8) | 0 (0.0) | NS |
| Cough (%) | 33 (73.3) | 59 (54.6) | 6 (35.3) | 15 (42.9) | 0.012 |
| Fever (%) | 33 (73.3) | 78 (72.2) | 12 (70.6) | 16 (45.7) | 0.024 |
| Sore throat (%) | 5 (11.1) | 18 (16.7) | 4 (23.5) | 10 (28.6) | NS |
| Asthenia (%) | 28 (62.2) | 41 (38.0) | 6 (35.3) | 11 (31.4) | 0.017 |
| Headache (%) | 9 (20.0) | 19 (17.6) | 3 (17.6) | 7 (20.0) | NS |
| GI symptoms (%) | 9 (20.0) | 15 (13.9) | 4 (23.5) | 1 (2.9) | NS |
| Dyspnea (%) | 1 (2.2) | 5 (4.6) | 0 (0.0) | 1 (2.9) | NS |
| Myalgia (%) | 14 (31.1) | 36 (33.3) | 9 (52.9) | 6 (17.1) | NS |
| Number of symptoms per patient (median, IQR) | 3 (2-4) | 2 (1-3) | 2 (2-4) | 2 (1-3) | NS |
| **Viral variant** |  |  |  |  |  |
| B.1.1.7/Alpha (%) | 40 (19.5) | 91 (44.4) | 15 (7.3) | 0 (0.0) | - |
| Q.4/Alpha (%) | 3 (1.5) | 12 (5.6) | 0 (0.0) | 0 (0.0) | - |
| BA.1/Omicron (%) | 0 (0.0) | 0 (0.0) | 0 (0.0) | 13 (38.2) | - |
| BA.1+R346K/Omicron (%) | 0 (0.0) | 0 (0.0) | 0 (0.0) | 14 (40.0) | - |
| BA.2/Omicron (%) | 0 (0.0) | 0 (0.0) | 0 (0.0) | 7 (20.0) | - |
| B.1.462 (%) | 1 (2.2) | 1 (0.9) | 0 (0.0) | 0 (0.0) | - |
| C.36.3 (%) | 0 (0.0) | 0 (0.0) | 1 (5.9) | 0 (0.0) | - |

SARS-CoV-2 whole-genome sequencing revealed variants belonging to five distinct clades, of which the most frequent were 20I/Alpha (n = 161), 21K/Omicron (n = 27), and 21L/Omicron (n = 7). Patients receiving bamlanivimab, bamlanivimab/etesevimab, or casirivimab/imdevimab mostly carried Alpha sub-variants (B.1.1.7, 146/170; Q.4, 15/170) at baseline except for 3 patients who carried 20A/B.1.462 or 20D/C.36.3 (Table 1). All patients treated with sotrovimab carried Omicron sub-variants, the most common being 21K/BA.1 with the S:R346K substitution (n = 14; BA.1.1, BA.1.1.1), followed by 21K/BA.1 (n = 13; BA.1, BA.1.17, BA.1.17.2), and 21L/BA.2 (n = 7; BA.2, BA.2.9).

Differences in viral load in patients undergoing different mAb treatments were longitudinally studied by comparing cyclic threshold (Ct) values for open reading-frame (ORF1)ab-, N protein-, and S protein-encoding genes by RT-qPCRs. A gradual, significant increase in cyclic threshold (Ct) values was observed for all gene targets indicating a decreasing viral load (**Figure 1A, Supplementary Table 2–3**). Due to the S:Δ69/70 deletion in Alpha (B.1.1.7, Q.4) and BA.1(+R346K)/Omicron sub-variants, most samples were qPCR-negative for the *S* gene. Compared to patients infected with Alpha sub-variants, patients carrying Omicron sub-variants showed significantly higher viral loads before mAb infusion (D0) that stayed significantly higher till 48h after mAb infusion (D2 timepoint) (**Figure 1B**).

**Figure 1.**
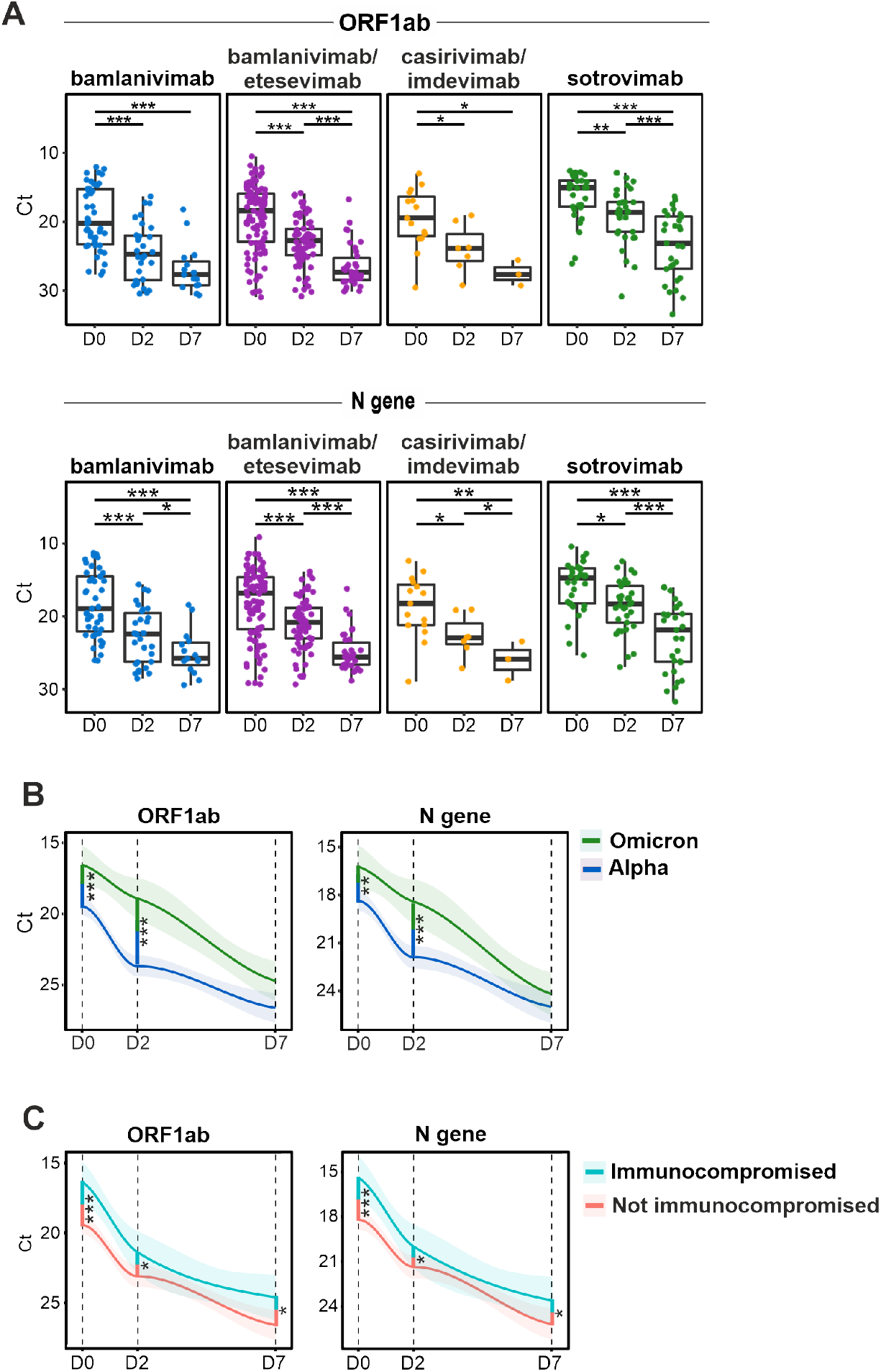
Immunocompromised and Omicron-infected COVID-19 patients display higher viral loads after mAb administration. Quantitative real-time reverse transcription (RT-q)PCR detection of SARS-CoV-2 was performed on nasopharyngeal swab (NPS) samples collected at D0, D2, and D7 from patients treated with different therapeutic mAbs. **(A)** A steady increase in Cyclic threshold (Ct) values was observed over 7 days for all mAb-treated groups. **(B)** Overall, patients carrying Omicron (BA.1, BA1+R346K, or BA.2) displayed higher viral loads than patients carrying Alpha sub-variants (B.1.1.7 or Q4). **(C)** Immunocompromised patients carry higher viral loads, irrespective of the infecting SARS-CoV-2 variant and mAb treatment. mAb: monoclonal antibody. D0: sample collected prior to mAb infusion. D2: 2 ± 1 days after mAb infusion. D7: 7 ± 2 days after mAb infusion. D28: 28 ± 4 days after mAb infusion.*: *p* < 0.05. **: *p* < 0.01. ***: *p* < 0.001. A limited number of NPS samples were collected at day 28 (n = 9) across all 4 mAb therapy groups and were therefore excluded from this analysis. See **Supplementary Table 2** and **3** for details on Ct values at each timepoint.

As several studies have shown that immunocompromised individuals show a prolonged carriage of SARS-CoV-2 (5, 7), we investigated whether these patients receiving mAb therapy also carried higher viral loads. Immunocompromised status was defined clinically on the basis of patients on active immunosuppressive treatment for cancer or immunological diseases, as described (19, 20). We showed that immunocompromised patients had higher viral loads at the time of enrolment within the mAb treatment groups (ΔCt 3.03 and 2.76 for *ORF1ab* and *N*, respectively; *p* < 0.001). Remarkably, significantly higher viral loads persisted in immunocompromised patients at both D2 and D7 timepoints (ΔCt at D7, 1.89 and 1.79 for *ORF1ab* and *N*, respectively; *p* ≤ 0.03) (**Figure 1C**). These data suggest that prolonged viral shedding occurs in immunocompromised COVID-19 patients with mild-to-moderate disease despite receiving mAb therapies.

### Immunocompromised patients display higher rates of SARS-CoV-2 Spike RBD mutations

To study the emergence of amino acid-substituting variants in response to mAb treatment, 204 patients were studied longitudinally for mutations occurring at D2 or D7 compared to pre-therapy (D0) timepoint. Forty-eight non-synonymous mutations at unique positions in the SARS-CoV-2 genome resulting in 43 amino acid substitutions were observed, of which 26/48 occurred spuriously (distributed across ORFs 1ab, 3a, and 7ab, or the *M* and *N* genes) and were observed only once (**Supplementary Figure 1, Supplementary Table 4**). The remaining 22 non-synonymous mutations occurred within the *S* gene, 16 of which occurred in Spike RBD (residues 319-541). As (RT-q)PCR errors have been suggested to be amplified to high allele frequencies resulting in sequencing errors, especially under low viral load conditions (8, 11), all non-synonymous Spike RBD mutations in sotrovimab-patients were re-confirmed by either Sanger or repeated NextSeq sequencing on independently extracted RNA.

A remarkable mutational homogeneity was identified wherein the same mutations developed independently in SARS-CoV-2 Spike RBD in different patients under mAb pressure. For instance, all 8 patients developing Spike RBD mutations receiving bamlanivimab or bamlanivimab/etesevimab involved only 3 residues (E484, Q493, S494, **Figure 2A-B; Supplementary Figure 2)**. Amongst these, Q493R was present in 3 patients and involved a residue common to both bamlanivimab and etesevimab binding sites suggesting a potential loss-of-function of binding of both mAbs to the SARS-CoV-2 mutated Spike protein. Similarly, mutations identified in sotrovimab-treated patients were present in either ACE2 (N417) or sotrovimab binding sites (D339, E340, R346, K440), except for 3 mutations involving residues L371, P373, and F375 identified in 3 patients (Figure 2B). These mutations involved alternate residues of SARS-CoV-2 Spike RBD and were notably substituted to serine, consistent with the Wuhan protein sequence. Two additional reversions (D339G and K346R) were identified in the sotrovimab-treated group, the latter mutation reversing the BA.1.1-defining R346K substitution (34).

**Figure 2.**
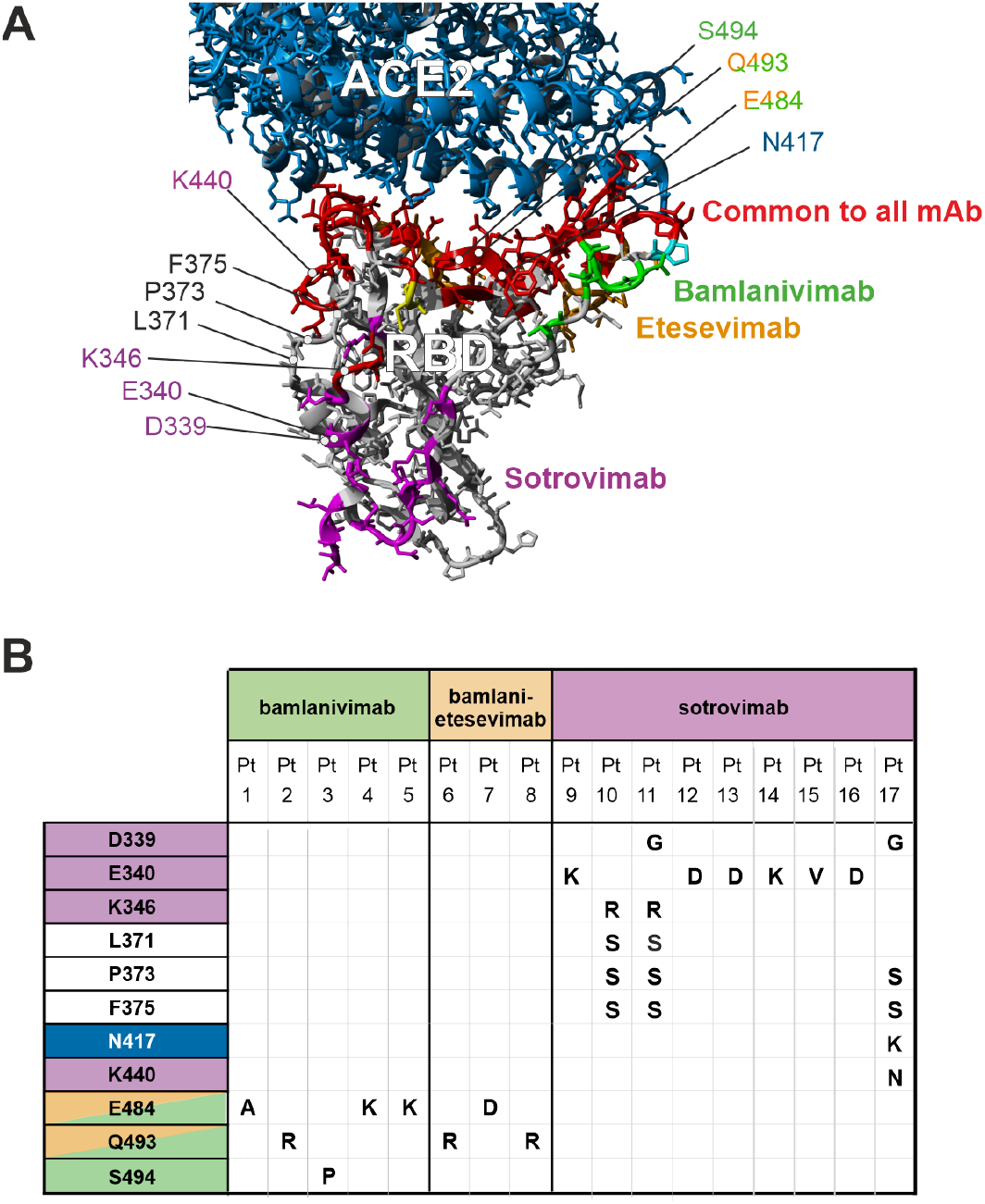
*De novo* SARS-CoV-2 Spike RBD mutations evolving under mAb pressure. **(A)** Schematic quarternary structure of the SARS-CoV-2 Spike RBD protein when bound to the human (h)ACE2 receptor (PDB: 6M0J). Key RBD-binding sites of bamlanivimab, etesevimab, and sotrovimab are highlighted in the protein structure with correpsonding colours. Binding sites common to all mAbs are indicated in red whereas hACE2 is highlighted in blue. **(B)** SARS-CoV-2 genomes longitudinally isolated from patients receiving mAb therapy were screened for the emergence of *de novo* mutations resulting in amino acid substitutions in the Spike RBD region. Most commonly, escape mutants ocurred in residues harbored within the respective mAb binding site. Pt: patient. For more details on non-synonymous *de novo* changes, see **Supplementary Figure 1** and **Supplementary Table 4**.

Notably, a highly diverse *S* gene mutation rate was also observed under the different mAb treatment:sub-variant combinations. For example, 9/34 (26.5%) patients carrying Omicron and receiving sotrovimab developed Spike RBD mutations, which was significantly higher compared to patients receiving other mAb treatments and carrying Alpha or other variants, i.e., 5/45 (11%) patients receiving bamlanivimab, 3/108 (2%) receiving bamlanivimab/etesevimab, and none (0/17) in the casirivimab/imdevimab group (Pearson χ^2^ = 21.005; n = 204; df = 3; *p* < 0.001).

Interestingly, patients with *de novo* mutations had approximately 10-fold increased quantity of viral genetic material at D0, D2, and D7 timepoints compared to patients without SARS-CoV-2 mutations across all mAb treatment groups (average ΔCt for *ORF1ab* and *N 3*.*37*, range 2.9–3.8, *p* ≤ 0.001). These data suggest that higher viral load predisposes to development of SARS-CoV-2 mutations. As immunocompromised individuals carried higher viral loads, we further assessed whether these individuals are more likely to develop Spike RBD mutations. Using a Chi-square test of independence, we showed a significant association between mAb treatments and development of mutations in the SARS-CoV-2 Spike RBD region in this population (Pearson χ^2^ = 4.633; n = 204; df = 1; *p* = 0.031). Together, these data suggest that COVID-19 patients receiving mAb therapy develop Spike RBD mutations that are not only mAb-therapy- or variant-dependent, but intra-host Spike mutations are also significantly increased in patients who are immunocompromised.

### Therapeutic mAb titers are not associated with development of Spike RBD mutations

We investigated anti-S and anti-RBD titers for different mAb treatment groups along with naturally developing anti-N titers at all timepoints. As sotrovimab was given to patients that were vaccinated (73.5%; 14 days post dose, ≥ 2 doses; n = 25; see Table 1), we first showed that, as expected, vaccine-related anti-S and anti-RBD titers, but not anti-N titers, were significantly elevated in the sotrovimab group at time of enrolment (D0) (**Supplementary Figure 3**). To address whether intervention with mAbs targeting SARS-CoV-2 could dampen the development of natural immunity, we studied anti-N titers that are not expected to be affected by therapeutic mAbs. A significant rise in anti-N titers was observed for all treatment groups, although the increase from pre-infusion titers (D0) to titers at D7 and D28 was smaller for the casirivimab/imdevimab and sotrovimab therapy groups compared to all others (**Figure 3A, Supplementary Table 5**). No significant difference in anti-S and anti-RBD titers was identified between patients infected with dominant circulating SARS-CoV-2 variants, including Omicron sub-variants (**Figure 3B**, Supplementary Table 5).

**Figure 3.**
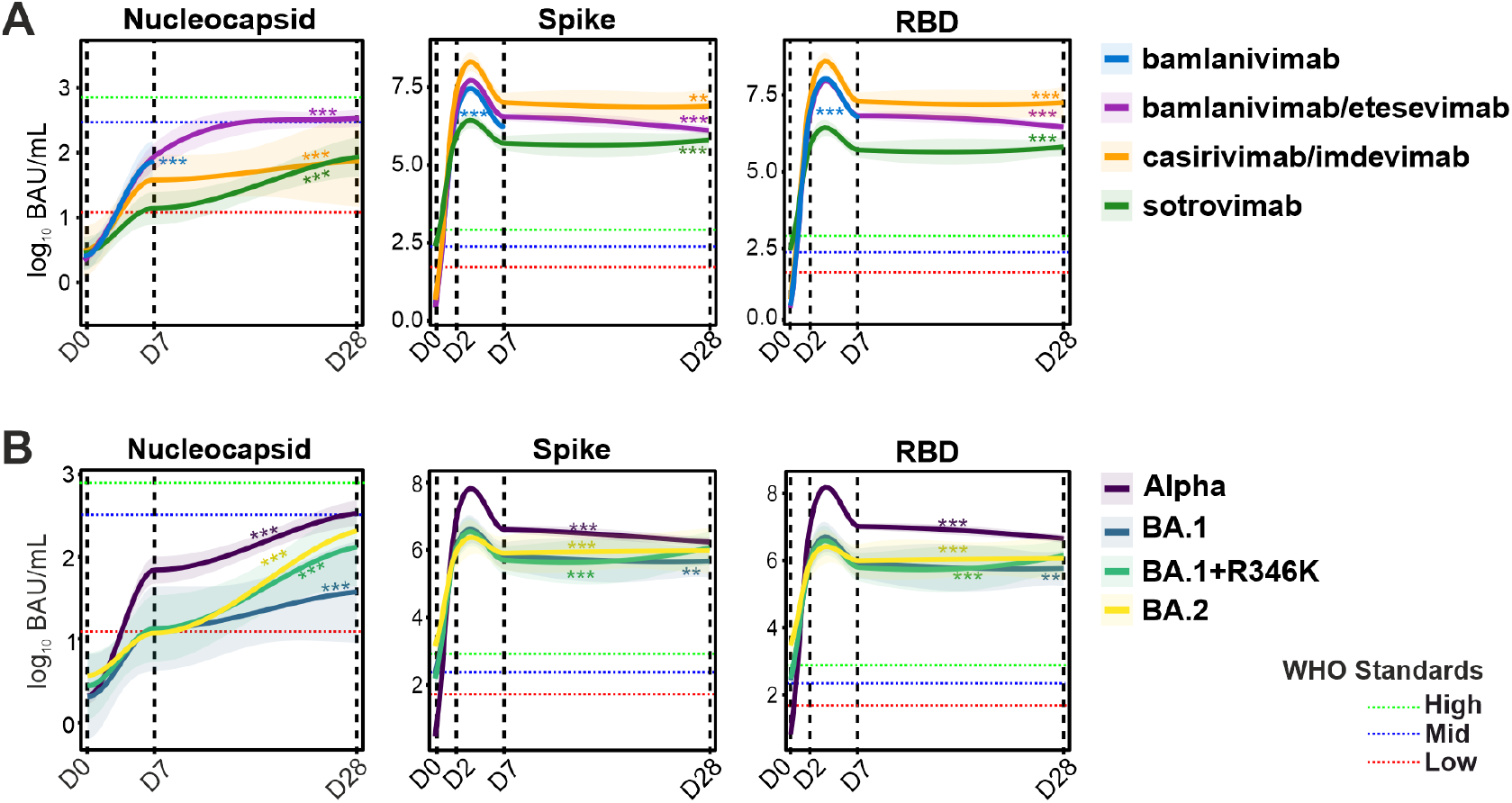
Temporal evolution of anti-N, anti-S, and anti-RBD serology titers in patients receiving mAb therapies. **(A)** Natural immunity was assessed based on anti-N titers, revealing a gradual increase through D28. High anti-S and anti-RBD titers due to therapeutic mAb administration persisted from D2 to D28 in patients in all treatment groups. **(B)** Similarly, high anti-S and anti-RBD titers were observed in patients receiving sotrovimab monotherapy carrying Omicron sub-variants (BA.1, BA1+R346K, or BA.2). Red, green, and blue dotted lines indicate SARS-CoV-2 WHO reference standard values for low, medium, and high antibody titers, respectively. *: *p* < 0.05. **: *p* < 0.01. ***: *p* < 0.001. Asterisks indicate significant slopes of the curves. For more details on serology in patients with or without vaccination, see **Supplementary Table 5**.

To study whether therapeutic antibodies could be linked to development of SARS-CoV-2 Spike RBD mutations, we first showed that pre-therapy (D0), the anti-S or anti-RBD titers were not significantly different in Spike RBD mutation carriers (n = 204; anti-S, F = 0.032, *p* = 0.859; anti-RBD, F = 0.140, *p* = 0.708). Similarly, we studied whether levels of therapeutic mAbs could be associated with Spike RBD mutations in our cohort. At the first post-therapy timepoint (D2), the average titers for anti-RBD and anti-S were 11.5 and 6.4 million BAU/mL, respectively. By comparison, the WHO International Standard of “High” corresponds to the anti-RBD titer of 817 BAU/mL and anti-S titers of 832 BAU/mL. Both anti-S and anti-RBD titers dropped at D7 and further at D28 for the majority of the mAb treatment groups, but the average anti-RBD and anti-S titers at D28 remained at 5.8 and 2.9 million BAU/mL, respectively. No association was observed between anti-S and anti-RBD titers at D2 or D7 and development of Spike RBD mutations. These data suggest that therapeutic mAb titers are exuberantly high in all patients and do not directly contribute to the development of SARS-CoV-2 Spike RBD mutations, although the sustained passive immunity could create an environment suitable for selection and sustenance of escape mutants.

### Neutralizing capacities of mAbs are (co-)drivers of development of escape mutants

While we show that therapeutic mAb titers *per se* are not related to development of mutations, we investigated whether the development of Spike RBD mutations is linked to the neutralization potential of different mAbs. Studying neutralizing capacities of the four mAb regimens in an ACE2 neutralization assay, we first showed a highly significant difference by which these mAbs neutralize five past or currently circulating SARS-CoV-2 variants (**Figure 4; Supplementary Figure 4**). Casirivimab/imdevimab appeared to have the highest neutralizing activity against most variants, including Wuhan, Alpha, and Omicron/BA.2 sub-variants. Sotrovimab monotherapy showed the best neutralization results against Omicron BA.1 (including BA.1+R346K sub-lineages), however, neutralizing activity against BA.2 was poor.

**Figure 4.**
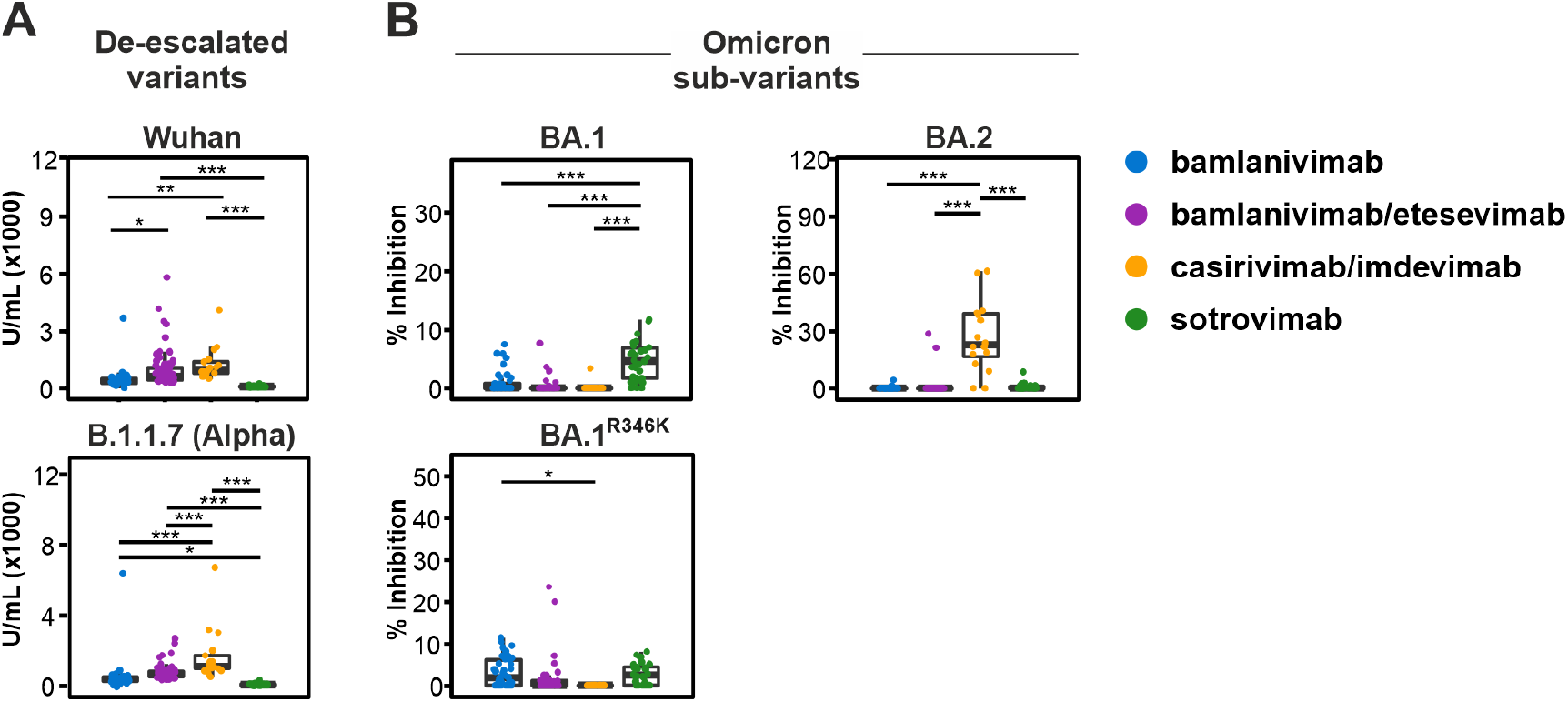
Anti-S neutralization capacity of bamlanivimab, bamlanivimab/etesevimab, casirivimab/imdevimab, and sotrovimab was measured against (A) de-escalated variants and (B) Omicron sub-variants at D2. Sotrovimab monotherapy proved most effective in neutralizing BA.1 sub-variants. Bamlanivimab showed increased neutralizing activity against BA.1 sub-variants. Casirivimab/imdevimab combination therapy proved highly effective in neutralization of BA.2 sub-variants. *: *p* < 0.05. **: *p* < 0.01. ***: *p* < 0.001. For details on variants of concern tested here, see **Supplementary Table 6**.

Remarkably, in the sotrovimab-treated group, both BA.1 and BA.2 infections were observed allowing us to assess whether neutralizing potential of mAbs could increase the likelihood of development of Spike RBD mutations. We show that for BA.1 and BA.1+R346K groups against which sotrovimab shows good neutralizing capacity, 9/27 (33%) of patients developed mutations. On the other hand, none of the patients in the BA.2 group (0/7), against which sotrovimab shows poor neutralizing capacity, developed Spike RBD mutations. These data were also statistically significant (likelihood ratio = 4.97, n = 34; df = 1, *p* = 0.026). Importantly, a higher proportion of immunocompromised patients (4/7, 57%) were present in the BA.2 group that did not develop mutations compared to the BA.1 group (13/27, 48%) (Spearman correlation, co-variance = 0.201, *p* = 0.708). These data strongly suggest that seroneutralization capacities of therapeutic mAbs are independently linked with development of SARS-CoV-2 escape mutants.

### Natural adaptive T-cell immunity facilitates development of SARS-CoV-2 escape mutants

Existing immunity against SARS-CoV-2 infection as a result of current or past exposure, vaccination against SARS-CoV-2, or human immune system variations could strongly influence the disease outcome in patients receiving different mAb regimens. To address the impact of mAb therapies on T helper (Th) cell immunity, lymphocytes collected at D0 and D28 were stimulated by either a SARS-CoV-2 Nucleocapsid or a complete Spike protein peptide pool (see Supplementary Methods). CD4^+^ Th cell-activation was subsequently studied by both a general marker, CD154 (CD40L), and by IFN-γ, a cytokine-associated marker of antigen-reactive Th cells.

At D0, the number of both S- and N-activated Th cells was significantly higher in the sotrovimab-treated group (n = 25) compared to bamlanivimab/etesevimab (n = 42) and casirivimab/imdevimab (n = 5) groups (*p* < 0.01) (**Figure 5A**). While the higher number of S-activated Th cells in the sotrovimab patients could be explained by vaccination, with most of the patients in this group being fully vaccinated, a concurrently higher number of N-activated Th cells suggests that patients in this group likely had prior exposure to SARS-CoV-2, as vaccination was administered to patients in this group later in the pandemic.

**Figure 5.**
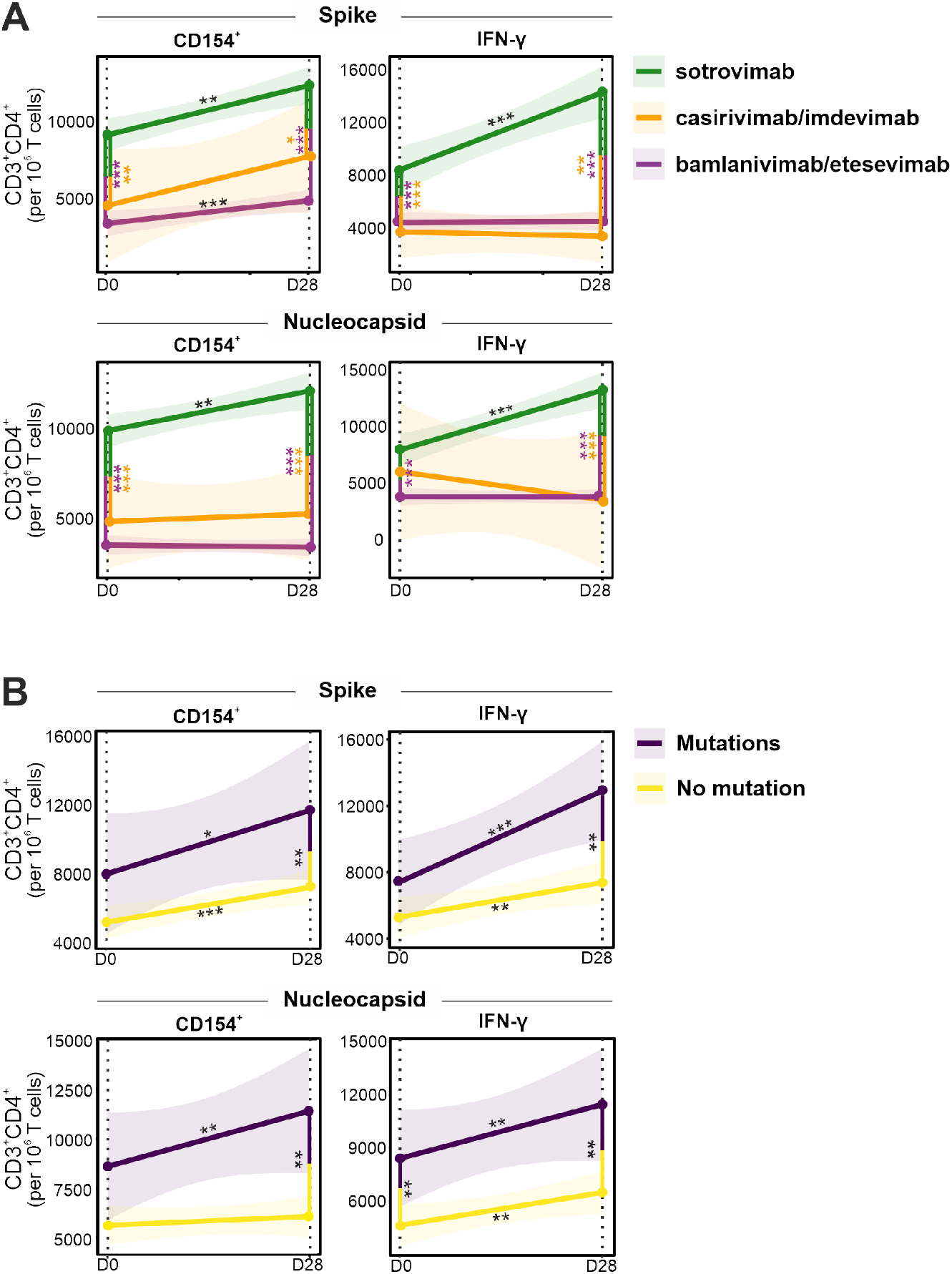
Longitudinal T cell responses in patients receiving mAb therapy. Evolution of IFN-γ and CD154 expression in SARS-CoV-2 Spike- and Nucleocapsid-stimulated CD4^+^ T cells in patients was studied over 28 days after receiving bamlanivimab/etesevimab, casirivimab/imdevimab, or sotrovimab. **(A)** Patients receiving sotrovimab therapy show a consistent significant increase in T cell expression during the first 28 days post mAb administration. For the utilized gating strategy, refer to **Supplementary Figure 7. (B)** Patients with *de novo* mutations in the SARS-CoV-2 Spike RBD region show an increased T cell expression compared to those without. Asterisks on regressions represent the significance of the slopes. Vertical lines with asterisks represent the significance of pairwise comparisons between patients with or without *de novo* mutations before mAb treatment (D0) and after 28 days of treatment (D28). *: *p* < 0.05, **: *p* < 0.01. ***: *p* < 0.001.

Furthermore, over 28 days, the sotrovimab-treated group also showed a significantly higher increase in N-activated CD4^+^IFN-γ^+^ cells compared to bamlanivimab/etesevimab and casirivimab/imdevimab groups (*p* < 0.001). These data suggest that sotrovimab does not majorly curb development of natural immunity and fits well with the significantly lower viral clearance observed in the sotrovimab-treated group carrying Omicron sub-variants compared to bamlanivimab/etesevimab and casirivimab/imdevimab groups carrying Alpha sub-variants (Figure 5A, Figure 1B).

Addressing whether mAb-induced Spike RBD mutations were associated with Th cell immunity, we further showed that patients developing mutations had slightly higher proportions of N-activated CD4^+^CD154^+^ and CD4^+^IFN-γ^+^ cells before mAb treatment, which was statistically significant for CD4^+^IFN-γ^+^ cells (*p* < 0.05; **Figure 5B**). However, strikingly, patients exhibiting *de novo* mutations also developed stronger Th cell immunity over 28 days with significantly increased S- and N-activated CD4^+^CD154^+^ and CD4^+^IFN-γ^+^ cells at 28 days (*p* < 0.01). While activated CD4^+^ Th cells could stimulate naïve B cells to produce specific antibodies against the mutant virus, these data strongly support the premise that the identified *de novo* mutations in the SARS-CoV-2 Spike protein are indeed escape mutations that evade therapeutic mAb neutralization, thereby facilitating a more natural progression of disease, resulting in more robust SARS-CoV-2–specific Th cell immunity.

### Host immune profile as a predictor of Spike RBD escape mutants

Studies have shown that pro-inflammatory cytokines when uncontrolled and exaggerated can lead to immunopathogenesis such as cytokine release syndrome disorder, however, under homeostatic conditions they are believed to play a major role in the control and resolution of SARS-CoV-2 infection (35, 36). Moreover, cytokines along with growth factors are critical to fundamental homeostatic processes such as wound healing and tissue repair (37). We hypothesized that a host environment that is, one, less hostile to the virus and, two, facilitates tissue repair, would together allow boosted cell infection cycles for rapid viral evolution under mAb pressure. To address this hypothesis, we studied 40 blood cytokines, chemokines, and growth factors as part of circulating immune-related biomarkers (CIB) involved in either COVID-19 pathogenesis and/or wound healing.

Significant alterations occurred in levels of 34/40 (85.0%) cytokines between different treatment groups (**Supplementary Figure 5**), which is also linked to infection with different SARS-CoV-2 variants. We further utilized area under the curve receiver operating characteristic (AUROC) analysis to discriminate between patients developing *de novo* Spike RBD mutations from those who did not or those who rapidly cleared the virus. AUROC for CIBs just before mAb administration identified 11 biomarkers to be significantly altered. Amongst these, 8 biomarkers were significantly increased in patients developing mutations and included angiogenic growth factors (bFGF, PlGF, and VEGF-D), angiogenic factors’ receptors (Tie-2 and Flt-1), and drivers of healing responses through macrophages (MCP-2 and MCP-3)(38) (**Figure 6A**). The four biomarkers that were significantly downregulated were acute phase inflammatory marker SAA, neutrophil chemokine IL-8, immunomodulatory marker IL-10, as well as M-CSF, a key cytokine involved in macrophage differentiation that enhances the inflammatory response of primed macrophages (39). Interestingly, after 48h of mAb infusion, the only cytokines observed to be significantly altered (n = 8) were those that were also significantly altered at D0 (**Figure 6B**). By day 7, several of these mutation-associated cytokines stayed altered (**Figure 6C**). These data suggest that, firstly, therapeutic mAbs do not majorly alter cytokine profiles in mildly ill COVID-19 patients, and secondly, cytokines identified to be linked to *de novo* Spike RBD mutation development are quite robust.

**Figure 6.**
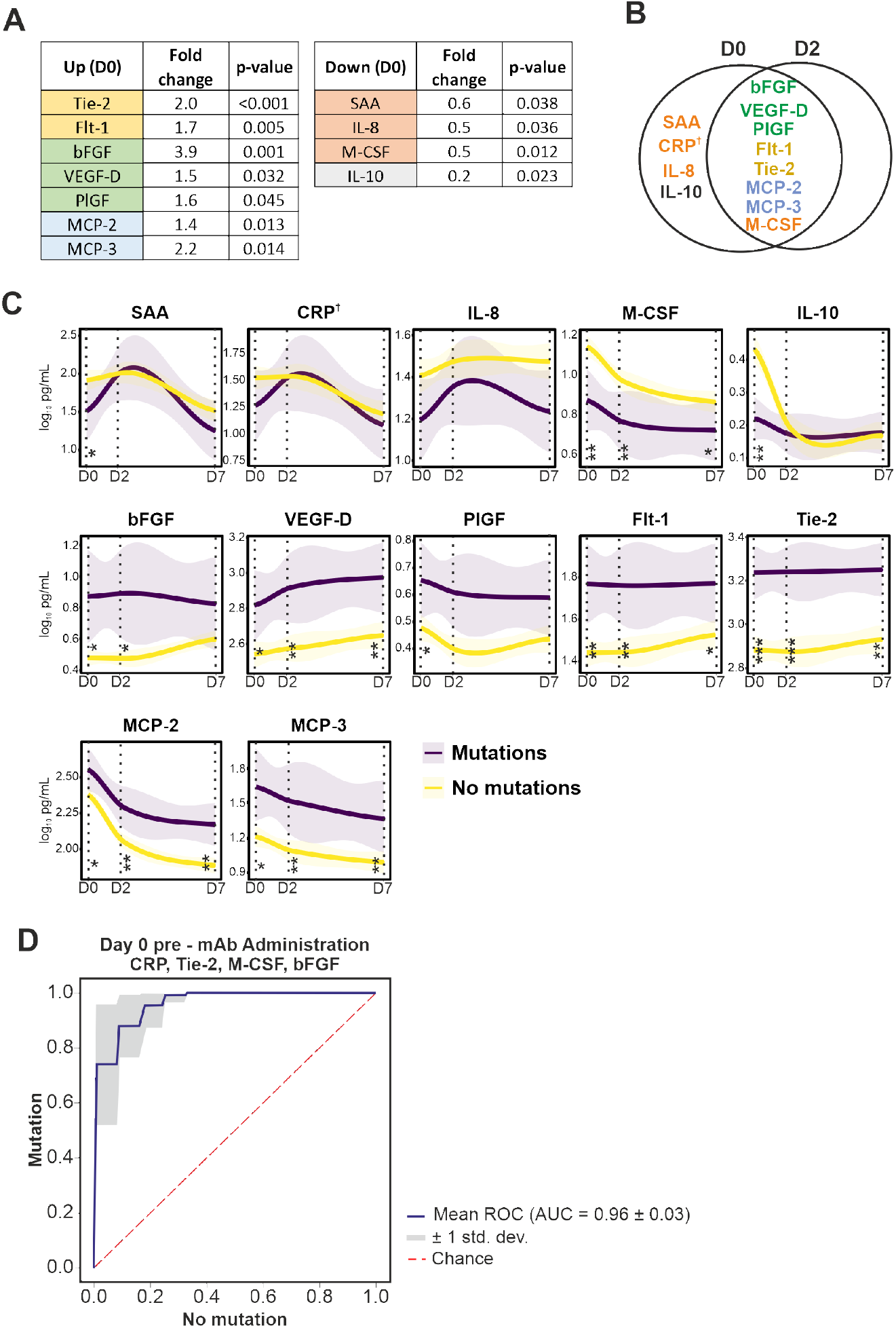
Circulating immune-related biomarkers (CIB) in COVD-19 patients receiving mAb therapy. **(A)** Several CIBs are significantly up- or downregulated at D0 in COVID-19 patients that developed SARS-CoV-2 Spike RBD mutations after administration of mAb treatments. **(B)** Eleven CIBs were significantly altered at D0 in patients with *de novo* Spike RBD mutations, for which the majority (n = 8) were also altered at D2. **(C)** Temporal evolution of CIBs altered in patients with or without *de novo* mutations, receiving mAb therapy through day 7 after treatment. P-values refer to significance of the slope of the regression lines. Vertical lines with asterisks represent the significant difference between CIB levels at the specified timepoints. **(D)** Receiving operator characteristic (ROC) curve in a random forest classifier model with Synthetic Minority Oversampling Technique (SMOTE) for the prediction of the qualitative response (mutation or no-mutation) are depicted for D0. *: *p* < 0.05. **: *p* < 0.01. ***: *p* < 0.001. †: not significant. For details on the progression of CIBs from D0 to D7, see **Supplementary Figure 5**.

AUROC data were further validated with Random Forest classification, which identified a signature consisting of SAA, Tie-2, bFGF, and M-CSF that correctly identified patients with *de novo* Spike RBD mutations with 96% accuracy. While CRP on its own missed statistical significance with AUROC analysis, replacing CRP with SAA did not change the accuracy of the model, likely because of high degree of co-linearity identified between CRP and SAA (Pearson’s R = 0.937, *p* < 0.001; **Figure 6D**). These data not only suggest that a diminished pro-inflammatory and homeostatic cytokine immune milieu could facilitate development of *de novo* Spike RBD mutations, but also describe a CIB profile present before mAb administration that predicts development of escape mutations against therapeutic mAbs for SARS-CoV-2 in high-risk patients with high accuracy.

## Discussion

Absence of virus from respiratory tract samples is suggested to occur once serum neutralizing antibody titers of 1:80 or 2,000 BAU/mL are achieved (32, 40). Considering that the average serum antibody titers in mAb-treated patients are more than a million BAU/mL, or 1000-fold higher than “high seropositivity” as defined by the WHO international standards, our data suggest that therapeutic mAbs are unable to readily cross the respiratory mucosal barrier and neutralize SARS-CoV-2. All therapeutic mAbs investigated in this study are IgG subtypes, and while special mechanisms such as receptor-mediated IgG transport exist, most of the mucosal humoral immunity is either mediated by IgA or extravasated plasma cells that then locally secrete immunoglobins including IgG (41-43). These data suggest that while therapeutic neutralizing mAbs efficiently clear SARS-CoV-2 from systemic tissue and reduce the risk of severe disease, the virus continues to thrive in the epithelial cells and mucosal barrier of the respiratory tract. With immunocompromised individuals exhibiting 4-fold higher viral loads compared to immunocompetent COVID-19 patients, these data not only support the evidence that immunocompromised patients have prolonged SARS-CoV-2 shedding (5, 7), but also suggest that innate cellular immunity is decisively involved in SARS-CoV-2 clearance from the respiratory tract. Our study design where patients received exogenous immunoglobins without affecting host plasma cells, also offers novel insights into the relative higher importance of local secretion of immunoglobins by mucosal plasma cells, as opposed to transepithelial transport of immunoglobins, in conferring mucosal immunity. These data can also be extrapolated to humoral mucosal immunity against other respiratory viral and bacterial pathogens.

Not only do we show that respiratory viral carriage is more abundant in immunocompromised patients, we also show that occurrence of *de novo* mutations is significantly higher in these patients, as shown for severely or chronically ill immunocompromised COVID-19 patients (8-12). Most mutations in SARS-CoV-2 are either deleterious or relatively neutral and only a small proportion impact virus characteristics including its transmissibility, virulence, and/or resistance to existing host immunity (4, 44). Concerns have also been raised that mutation rates could be overestimated due to reverse transcriptase or sequencing errors (11, 45). However, for the following reasons we believe that the identified mutations in Spike RBD are existent and non-neutral. First, the mutations were reconfirmed by performing sequencing on independently extracted RNA making it a high-fidelity observation. Second, Spike RBD mutations were identified 2-7 days after mAb treatment in contrast to studies where mutations were observed before treatment, for example, case studies where mutations in Spike were fixed before casirivimab/imdevimab treatment (8, 46). Third, the observed *de novo* mutations are highly specific to cognate mAb or ACE2 binding sites. For example, Spike RBD mutations developing in bamlanivimab- or bamlanivimab/etesevimab-treated patients had no overlap with mutated sites observed in sotrovimab-treated patients. Fourth, the *de novo* mutations are also highly evasive to therapeutic antibodies. For example, sotrovimab given empirically to BA2-infected patients, against which sotrovimab shows little neutralization, did not develop escape mutations, while it did for BA1-infected patients against which sotrovimab is highly active. Fifth, sotrovimab-receiving BA.1-infected patients had more robust SARS-CoV-2–specific Th cell immunity, likely due to lack of SARS-CoV-2 neutralization. And, lastly, possible non-neutrality of some mutations described in the study are supported by prior reports on identical or similar mutations (24-30) (see Supplementary Table 4). Amino acid residues typically observed in Omicron sub-variants reverting back to those of the original Wuhan sequence (D339G, L371S, P373S, F375S, N417K, and K440N) are equally interesting, some of which have also been observed previously (17), supporting our seroneutralization data showing that sotrovimab is not active against Wuhan and some of the other de-escalated variants.

While we show that the *de novo* Spike RBD mutations are unequivocally mAb-specific mutations, we also show that the mutations accumulate in acutely infected patients and occur rather rapidly, within 7 days of treatment. Prior studies have proposed that selection pressure created during chronic or severe infections drives the emergence of SARS-CoV-2 mutations (8-12). Our data suggest that neither chronic nature of the disease, nor its severity are necessary for occurrence of mutations if immune pressure is profound and rapid, as that induced by synthetic neutralizing mAb therapy. Both RNA and DNA viruses are capable of generating *de novo* diversity in a short period of time while adapting to new hosts and environments (47). Although one thing common to both our and previous studies is that the mutation rate is significantly higher in immunocompromised patients (8-12), we also show that higher viral loads, regardless of the cause, are directly linked to Spike RBD mutations development.

We identify two specific components of host immunity that are associated with these mutations. Firstly, we demonstrate that downregulated pro-inflammatory cytokines are linked with higher rate of mutations, likely due to decreased viral clearance and more replication cycles giving the virus a higher chance to adapt evolutionarily. Cytokine immunity is an important component of host innate and adaptive immunity, and while examples exist where pro-inflammatory cytokines could be suppressed by viruses (48), the cytokine alterations associated with *de novo* mutations are likely driven by host-genetic susceptibilities to SARS-CoV-2 (49). Secondly, in a mutually non-exclusive independent mechanism, we also show that patients developing *de novo* mutations had stronger Th cell immunity following mAb treatment, suggesting a strong immune pressure on the virus to adapt (6). Additionally, we describe an upregulation of key host growth factors, such as angiogenic growth factors and their receptors, that could be a consequence of SARS-CoV-2-induced lung damage. However, because patients only had mild disease, we propose that a reparative milieu, likely also genetically driven, while facilitating a rapid recovery of patients, could also allow boosted cell infection cycles enabling the virus to adapt. Our pharmacokinetic studies further showed that levels of all mAbs were retained at more than one million BAU/mL over 4 weeks, suggesting a sustained longstanding environment wherein mutant SARS-CoV-2 could be sheltered and mutate further, posing threats for viral rebound infections and dissemination of novel mutants. It is hypothesized that almost all SARS-CoV-2 variants originated in immunocompromised chronic carriers (50). Our data therefore emphasize the need of optimized mitigation strategies in immunocompromised patients receiving mAb treatment to reduce the risk of spreading of virus to other high-risk patients in both a hospital and community setting.

Lastly, we suggest that assessment of CRP or SAA (general marker of inflammation), bFGF (angiogenic ligand), Tie-2 (angiogenic growth factor receptor), and M-CSF (pro-inflammatory and immune regulatory mediator) in high-risk patients with SARS-CoV-2 infection under evaluation for mAbs therapy could identify patients, with high accuracy, who are also at risk of developing escape mutations against therapeutic mAbs. This or similar biomarker-based stratification could also benefit decision making. For example, identification of immunocompromised patients who are also at high risk of developing *de novo* mutations could benefit from alternative strategies such as anti-viral treatments.

As limitations, samples analyzed in this study were collected during an extended time-period, resulting in underlying differences in the patient population, such as rate of vaccination and circulating SARS-CoV-2 variants. At the same time, the heterogeneity of infecting VOCs and inclusion of vaccinated individuals among high-risk groups could be considered a strength of the study, as this enables representation of real-world data and rapid changes in epidemiological scenarios typical of the SARS-CoV-2 pandemic. Being a prospective monocentric cohort within a European project, this study had the advantage of homogenous sampling and enrolment protocol but lacks external validity for cytokine-based biomarkers. Finally, a very limited number of nasopharyngeal swab samples were collected at D28, thereby not allowing us to study the impact of mutation on prolonged carriage.

Despite these limitations, we show in a comprehensive analysis of patients with diverse mAb treatments, development of adaptive mutations that highly correlate with neutralizing capacities of therapeutic mAbs and provide direct evidence that anti-SARS-CoV-2 host-driven responses are necessary and essential for development of mutant SARS-CoV-2. While these data, on one hand, suggest a critical balance between successful viral killing and development of VOC-like mutations in niched environments such as respiratory mucosa, on the other hand, our data also prompts close and extensive monitoring and isolation of patients and contacts to limit the spread of potential VOC-like mutants, especially in high-risk populations.

## Material and Methods

### Study design

Samples were collected as part of the prospective, observational, monocentric ORCHESTRA cohort study conducted from 9 March 2021 to 22 August 2022 in the early COVID-19 treatment Outpatient Clinic, Infectious Diseases Section of the University Hospital of Verona, Italy. All outpatients aged ≥ 18 years, presenting with mild-to-moderate COVID-19 (confirmed by quantitative real-time Reverse Transcription (RT-q)PCR or a positive antigenic 3^rd^ generation test) at high risk for clinical worsening in accordance with Italian Medicine Agency indications (for definition see ref. (19, 20)) were offered monoclonal antibody therapy and enrolled in this study. All enrolled patients received treatment with either bamlanivimab, bamlanivimab/etesevimab, casirivimab/imdevimab, or sotrovimab.

Further details on patient inclusion/exclusion criteria and other clinical criteria have recently been published (19, 20). Ethical approval for this study was obtained from the University Hospital Verona Ethics Board (protocol number: 19293) and conducted in accordance with the Declaration of Helsinki. No study activities took place prior to collection of informed consent.

Samples were collected from enrolled patients to study the effect of mAb therapy on SARS-CoV-2 viral load, mutations induced by different mAbs, mAb kinetics, neutralization capacity of mAb, cellular immunity, and cytokine responses. For each enrolled patient, 4 timepoints were analyzed: i) D0: just prior to mAb infusion, (ii) D2: 2 ± 1 days after mAb infusion at D0, (iii) D7: 7 ± 2 days after mAb infusion at D0, and (iv) D28: 28 ± 4 days after mAb infusion. Nasopharyngeal swab, serum, and Peripheral blood mononuclear cells (PBMCs) samples were collected along with clinical data. An overview of the number of samples included for each analysis is available in **Supplementary Figure 6**.

### SARS-CoV-2 viral load and variant sequencing

RNA was extracted using the MagMAX Viral/Pathogen II Nucleic acid kit on a KingFisher Flex Purification System (ThermoFisher, the Netherlands). Real-Time RT-qPCR was performed using the TaqPath™ COVID-19 CE-IVD RT-PCR Kit (ThermoFisher, the Netherlands) on a QuantStudio™ 5 Real Time PCR instrument (384-well block, 5 colors, ThermoFisher, the Netherlands). Extracted RNA was subjected to automated cDNA conversion and multiplexed library preparation using the Illumina COVIDSeq Test kit (Illumina Inc., CA, USA) on a Zephyr G3 NGS (PerkinElmer, MA, USA) and sequenced using the High Output Kit v2 on a NextSeq 500/550 instrument (Illumina Inc., CA, USA). Identified single nucleotide polymorphisms (SNPs) were verified by Sanger sequencing. For detailed methods, refer to Supplementary Information.

### Serology

Blood was collected in 10 mL serum tubes (BD vacutainer K2E, BD biosciences, Germany) and serum samples were prepared within 3h of blood collection. Anti-N, anti-S, and anti-RBD SARS-CoV-2 IgG titers were measured in serum samples using a multiplexed panel (Meso Scale Discovery (MSD), MD, USA). For detailed methods, refer to Supplementary Information.

### ACE2 neutralization measurements in serum

ACE2 neutralization measured in serum samples using V-PLEX SARS-CoV-2 Panel 6, Panel 13, Panel 23 and Panel 25 Kits (ACE2) on the QuickPlex SQ 120 system (MSD, MD, USA) according to the manufacturer instructions. Details regarding the Spike variants, against which the neutralizing antibody titers were measured, are displayed in **Supplementary Table 8**. For detailed methods, refer to Supplementary Information.

### Measurements of circulating immune-related biomarkers (CIB) in serum

CIBs were measured in serum samples using U-plex and V-plex panels (#K15198D, #K15190D) on the QuickPlex SQ 120 system (MSD, MD, USA), according to the manufacturer instructions. The following 40 CIBs were measured for D0, D2, and D7 timepoints: basic fibroblast growth factor (bFGF), C-reactive protein (CRP), cutaneous T-cell attracting chemokine (CTACK), eotaxin, erythropoietin (EPO), vascular endothelial growth factor receptor 1 (Flt-1), fractalkine, macrophage colony-stimulating factor (M-CSF), interferon β (IFN-β), interferon γ (IFN-γ), interleukin (IL)-1β, IL-1 receptor antagonist (IL-1Ra), IL-2, IL-2 receptor α (IL-2Rα), IL-4, IL-5, IL-6, IL-8, IL-10, IL-13, IL-15, IL-17A, IL-17F, IL-18, IL-22, IL-33, IFN-γ induced protein 10 (IP-10), monocyte chemoattractant protein (MCP)-1, MCP-2, MCP-3, macrophage inflammatory protein (MIP)-1α, placental growth factor (PlGF), serum amyloid A (SAA), soluble intercellular adhesion molecule 1 (sICAM-1), soluble vascular cell adhesion molecule 1 (sVCAM-1), angiopoietin receptor 1 (Tie-2), tumor necrosis factor α (TNF-α), vascular endothelial growth factor (VEGF)-A, VEGF-C, and VEGF-D. For detailed methods, refer to Supplementary Information.

### SARS-CoV-2 specific cellular responses

PBMCs were isolated using cellular preparation tubes (BD Biosciences, Germany) according to the manufacturer’s instructions and stored in fetal bovine serum (FBS) with 10% DMSO at -80°C until further use. Stimulation and staining were performed using the SARS-CoV-2 T Cell Analysis Kit (PBMC), human (Miltenyi Biotech, Germany). PBMCs were stimulated with a pool of lyophilized peptides, consisting of 15-mer sequences covering the complete protein-encoding sequence of the SARS-CoV-2 surface or Spike glycoprotein (GenBank MN908947.3, Protein QHD43416.1) and the complete sequence of the nucleocapsid phosphoprotein (GenBank MN908947.3, Protein QHD43423.2) from Miltenyi Biotech. For detailed methods, refer to Supplementary Information.

### Flow cytometry

After stimulation, staining of surface and intracellular antigens was carried out with the following fluorochrome-conjugated recombinant human IgG1 isotype antibodies (Miltenyi Biotech, Germany): CD3-APC REAfinity™ (clone REA613), CD4-Vio® Bright-B515 REAfinity (clone REA623), CD8-VioGreen™ REAfinity (clone REA734), CD14-CD20-VioBlue® REAfinity (clone REA599, clone REA780), IFN-γ-PE REAfinity (clone REA600), TNF-α-PE-Vio® 770 REAfinity (clone REA656), CD154-APCVio® 770 REAfinity (clone REA238). Samples were captured on a NovoCyte Quanteon 4025 flow cytometer (Agilent, CA, USA) and analyzed using FlowJo v10.8.1 (BD, NJ, USA) (**Supplementary Figure 7**). For detailed methods, refer to Supplementary Information.

### Statistical analysis

All data were statistically analyzed and visualized in Rstudio v.1.3.1073 using R v.4.0.4. One-way analysis of variance (ANOVA) was utilized for longitudinal and cross-sectional comparisons of IgG titers, titers of neutralizing antibodies, and CIB concentrations across treatment groups followed by pairwise two-tailed t-tests. Cyclic threshold (Ct) values were compared using non-parametric Kruskal-Wallis followed by pairwise testing using Mann-Whitney. Post hoc p-value correction was conducted using Bonferroni’s multiple-comparison correction method for all analyses. Throughout the statistical analyses, values below the detection range were recorded as 1/10^th^ the lower limit of quantitation (LLQ) and values above the detection range were recorded as upper limit of quantitation (ULQ). A (corrected) p-value < 0.05 was considered statistically significant. For the identification of the main predictors of qualitative responses (mutation/no mutation in the Spike RBD region [residues 319-541]), receiver operating characteristic (ROC) curves were constructed utilizing MetaboAnalyst. Machine-learning-based Random Forest classifiers (RFC) were further built by the Python package sklearn v2.0 to independently predict development of *de novo* Spike RBD mutations in patients receiving mAb regimens. Each model was built with a training set of values consisting of 80% of the data and a test set of 20%(51). To account for imbalanced groups, the Synthetic Minority Oversampling Technique (SMOTE, Python package imblearn 0.8.0) was utilized in combination with the RCF method. The models were bootstrapped 100 times and features for each model were selected based on i) feature importance, ii) statistics from mutation vs. non-mutation, iii) individual ROC curve analysis, and iv) a Pearson correlation matrix for independence of variables. Confusion matrices and ROC curves were drawn to calculate area under the curve (AUROC) values to verify reliability and to evaluate the performance of the constructed models. For data visualization, box plots indicate median (middle line), 25th, 75th percentile (box), and 5th and 95th percentile (whiskers). All data points, including outliers, are displayed. Line graphs display 95% confidence intervals for all measured timepoints.

## Data Availability

Data supporting the findings of this study are available within Supplementary Information files. SARS-CoV-2 genome sequences obtained in the project were submitted to GISAID. Trimmed read data generated and used for identification of emerging de novo Spike RBD mutants in this study have been submitted to the European Nucleotide Archive (ENA) under the project accession PRJEB55794. All other data generated in this study are available from the corresponding author upon request.

## Data availability

Data supporting the findings of this study are available within Supplementary Information files. SARS-CoV-2 genome sequences obtained in the project were submitted to GISAID. Trimmed read data generated and used for identification of emerging *de novo* Spike RBD mutants in this study have been submitted to the European Nucleotide Archive (ENA) under the project accession PRJEB55794. All other data generated in this study are available from the corresponding author upon request.

## Author Contribution

Conceptualization: SKS, ET, SMK, PDN; Overall study supervision: SKS; Clinical data and sample collection: AS, PDN, MM, DP, ED, ER, ET; RT-qPCR and viral variant sequencing: MS, MB, SMK; Bioinformatic analysis: MB, MS; Serological analysis: AK, AG, FDW; Cytokine, chemokine, and growth factor analysis: AK, AG, FDW, AH; PBMC isolations: DP, PD, ER; PBMC analysis: AG, VVA, AK, FDW; Statistical analysis: MB, AG, AK, FDW, SKS; Data interpretation: SKS, SMK, ET, AG, AK, MB, MS; Manuscript writing: SKS, MB, AG, AK, MS, SMK, ET; All authors read, gave input, and approved the final manuscript.

## Supplementary Information

### Supplementary Methods

#### RNA extraction, cDNA conversion, library preparation, and SARS-CoV-2 whole genome sequencing

RNA was extracted using the MagMAX Viral/Pathogen II Nucleic acid kit on a KingFisher Flex Purification System (ThermoFisher, the Netherlands). Each batch of samples taken forward for extraction was processed together with a Twist synthetic SARS-CoV-2 RNA positive Ctrl. 18 (Cat. No: 104338, Twist Bioscience). Extracted RNA was subjected to automated cDNA conversion and multiplexed library preparation using the Illumina COVIDSeq Test kit (Illumina Inc., CA, USA) on a Zephyr G3 NGS system (PerkinElmer, MA, USA). DNA concentrations were quantified using the Qubit dsDNA HS Assay Kit (Invitrogen, Cat. No. Q33231) using a Qubit Fluorometer 3.0 (ThermoFisher). Pooled libraries were sequenced utilizing the High Output Kit v2 with a 1.4 nM PhiX Library positive control v3 using a 1% spike-in on a NextSeq 500/550 instrument (Illumina Inc., CA, USA). All steps were performed according to manufacturer’s instructions.

#### SARS-CoV-2 RT-qPCR

Real-Time RT-qPCR was performed using the TaqPath™ COVID-19 CE-IVD RT-PCR Kit (ThermoFisher, the Netherlands) on a QuantStudio™ 5 Real Time PCR instrument (384-well block, 5 colors, ThermoFisher, the Netherlands), which detects three genes in the SARS-CoV-2-viral genome: the S protein, N protein, and ORF1ab. MS2 (phage control) was added to each sample prior to RNA extraction to serve as internal control. RT-qPCR analysis was performed using FastFinder (UgenTec, Belgium) according to the TaqPath™ COVID-19 CE-IVD RT-PCR Kit Ct cut-off values. Samples were considered positive if both the MS2 phage control (Ct < 32) and at least two gene targets were detected (Ct < 37).

#### SARS-CoV-2 variant detection

Raw sequencing data quality for each sample was assessed using FastQC (https://www.bioinformatics.babraham.ac.uk/projects/fastqc/) followed by quality trimming using a Phred score cut-off of 25 with TrimGalore v. 0.6.7 (https://github.com/FelixKrueger/TrimGalore). Read mapping was performed against the SARS-CoV-2 genome (GenBank: NC_045512.2) using the CLC Genomics Workbench v.9.5.3 (Qiagen, Germany) with a length and a similarity fraction of 0.5 and 0.8, respectively. Consensus sequences were extracted, and clade and lineage assignment performed using Nextstrain (https://clades.nextstrain.org/) and Pangolin (https://pangolin.cog-uk.io/), respectively. SARS-CoV-2 genome sequencing was considered successful if: i) was successfully classified by both Pangolin and NextClade, and ii) the resulting genome sequence harbored < 15% ambiguous base calls (Ns) in the consensus sequence.

For detection of single nucleotide variations (SNPs) acquired during monoclonal antibody treatment in patients who provided samples at D0, as well as D2 and/or D7, trimmed reads were mapped against the SARS-CoV-2 genome (GenBank: NC_045512.2) using the CLC Genomics Workbench v.9.5.3 (Qiagen, Germany) with a length and a similarity fraction of 0.7 and 0.99, respectively. SNPs resulting in amino acid substitutions were of particular interest and analyzed further in this study.

#### SNP validation using Sanger sequencing

Patients harboring multiple amino-acid-altering mutations in the Spike RBD region (residues 319 – 541) were subjected to Sanger sequencing to validate identified mutations. For this purpose, RNA extraction and cDNA conversion was repeated and used for the validation.

Primers binding in the region of interest were selected from the Artic primer pool v3 (nCoV-2019 sequencing protocol v3 (LoCost) by performing *in silico* PCR using the CLC Genomics Workbench v.9.5.3 (Qiagen, Germany) or designed using NCBI PrimerBlast with standard parameters (National Center for Biotechnology Information, MD, USA) utilizing the Wuhan (GenBank: NC_045512.2) and Omicron/BA1.1 (GenBank: OM664849) genomes as templates with the following criteria: i) optimal primer length = 25 bp, ii) >5 bp difference in length between forward and reverse primers, and iii) ΔTm <5°C. Designed primer pairs were validated *in silico* using FastPCR (https://primerdigital.com/) using standard parameters.

PCR amplification was performed using 50 ng cDNA, Q5 Hot start 2x MM (New England Biolabs, MA, USA), forward and reverse primers at a final concentration of 0.5 μM each, and Nuclease-free water (Ambion, ThermoFisher, the Netherlands) in a total volume of 45 μL with the following temperature profile: 98°C for 15s followed by 35 cycles of denaturation at 96°C for 30 s and annealing at 63°C for 5 min. Successful amplification was confirmed with 1.5% agarose gel electrophoresis (150 V, 200 mA, 1h) using a 100 bp DNA ladder (ThermoFisher, the Netherlands).

Obtained PCR products were then subjected to automatic template clean-up and sample preparation using Illustra™ ExoProStar™ (Merck, Germany) and ABI PRISM® BigDye™ Terminator cycle sequencing kits (ThermoFisher, the Netherlands) with Biomek® FX and NX liquid handlers (Tecan, Switzerland), followed by sequencing on an Applied Biosystems 3730XL DNA Analyzer (ThermoFisher, the Netherlands). Sequence analysis was performed using the CLC Genomics Workbench v.9.5.3 (Qiagen, Germany).

#### Serology

Blood was collected in 10 mL serum tubes (BD vacutainer K2E, BD Biosciences, Germany) and serum samples were prepared within 3h of blood collection. Serum was allowed to clot thoroughly for 60 min before separation by centrifuging for 10 min at 1300 g. Aliquots were flash frozen in liquid nitrogen, shipped to the University of Antwerp for further processing and stored at -80°C until analysis.

IgG titers were measured in serum samples using the V-PLEX SARS-CoV-2 Panel 6 Kit (IgG; #K15433U-4) on a QuickPlex SQ 120 instrument (Meso Scale Discovery (MSD), MD, USA) according to the manufacturer’s instructions. IgG titers to the following antigens were measured: SARS-CoV-2 Nucleocapsid, SARS-CoV-2 S1 RBD, SARS-CoV-2 Spike, SARS-CoV-2 Spike (D614G), SARS-CoV-2 Spike (B.1.1.7/Alpha), SARS-CoV-2 Spike (B.1.351/Beta), SARS-CoV-2 Spike (P.1/Gamma). Baseline samples were measured at 1:1,000 or 1:10,000, while all other samples were measured at a final dilution of 1:10,000,000 or 1:100,000,000 in Diluent 100 (MSD, MD, USA). Quantitative IgG results were measured in Antibody Units (AU)/mL converted to Binding Antibody Units (BAU)/mL using a conversion factor provided by the manufacturer and reported as such. Patients were considered negative if their levels were under 4.76 BAU/mL for anti-spike, under 5.58 for anti-RBD, and under 8.20 for anti-nucleocapsid, these limits were determined by calculating the average plus one standard deviation of IgG measurements in 56 serum samples collected before 2019.

#### ACE2 neutralization measurements in serum

ACE2 neutralization measured in diluted serum samples (1:3,000) using V-PLEX SARS-CoV-2 Panel 6, Panel 13, Panel 23 and Panel 25 Kits (ACE2) and measured on the QuickPlex SQ 120 instrument (MSD, MD, USA) according to the manufacturer’s instructions. Details regarding the Spike variants, against which the neutralizing antibody titers were measured, are displayed in **Supplementary Table 8**. Quantitative ACE2 neutralization results were measured in Units (U)/mL for all variants except Omicron sub-variants, which corresponds to neutralizing activity of 1 μg/mL monoclonal antibody to SARS CoV-2 Spike protein (upper limit of quantitation: 63,000 U/mL; lower limit of quantitation: 15 U/mL). Omicron sub-variants were measured as percent inhibition (% inhibition) calculated as 100 x [1 - (Sample signal/Average signal of the blanks)].

#### SARS-CoV-2 specific cellular responses

Peripheral blood mononuclear cells (PBMCs) were isolated using cellular preparation tubes (BD Biosciences, Germany) according to the manufacturer’s instructions and frozen in fetal bovine serum (FBS) with 10% DMSO until further use. Stimulation and staining were performed using the SARS-CoV-2 T Cell Analysis Kit (PBMC) human (Miltenyi Biotech, Germany). Briefly, PBMCs were thawed and rested overnight in RPMI 1640 medium (Gibco, ThermoFisher, the Netherlands) supplemented with 5% heat-inactivated AB serum (Sigma-Aldrich, Merck, Germany), 100 U/ml penicillin (Biochrom, Germany), and 0.1 mg/ml streptomycin (Biochrom, Germany). 1e6 PBMCs were stimulated with a pool of lyophilized peptides, consisting mainly of 15-mer sequences with 11 amino acids (aa) overlap, covering the complete protein coding sequence (aa 5–1273) of the SARS-CoV-2 surface or Spike glycoprotein of SARS-CoV-2 (GenBank MN908947.3, Protein QHD43416.1) and the complete sequence of the nucleocapsid phosphoprotein (GenBank MN908947.3, Protein QHD43423.2) from Miltenyi Biotech. Both peptide pools were used at 1 μg/mL per peptide. Stimulation controls were performed with equal concentrations of sterile water/10% DMSO (unstimulated) as negative control and Cytostim™ (Miltenyi Biotech, Germany) as positive control. Incubation was performed at 37 °C, 5% CO2 for 6h with 2 μg/mL brefeldin A (Sigma-Aldrich, Merck, Germany) added after 2 h.

#### Flow cytometry

After stimulation, staining of surface and intracellular antigens was carried out with the following fluorochrome-conjugated recombinant human IgG1 isotype antibodies (Miltenyi Biotech, Germany) at 0.25x recommended volume: CD3-APC REAfinity™ (clone REA613), CD4-Vio® Bright-B515 REAfinity (clone REA623), CD8-VioGreen™ REAfinity (clone REA734), CD14-CD20-VioBlue® REAfinity (clone REA599, clone REA780), IFN-γ-PE REAfinity (clone REA600), TNF-α-PE-Vio® 770 REAfinity (clone REA656), CD154-APCVio® 770 REAfinity (clone REA238). Cells were washed with cell staining buffer (PBS 1% bovine serum albumin, 2mM EDTA) unless stated otherwise. Briefly, dead cells were stained for 10 min with Viobility 405/452 Fixable Dye (1:200) with subsequent fixation and permeabilization for 20min (Inside stain kit, Miltenyi Biotech, Germany). Cells were washed with permeabilization buffer and surface marker, and intracellular staining was carried out for 15 min. Cells were washed in permeabilization buffer and resuspended in cell staining buffer. Samples were captured on a NovoCyte Quanteon 4025 flow cytometer (Agilent, CA, USA) and analyzed using FlowJo v10.8.1 (BD, NJ, USA) (**Supplementary Figure 7**).

### Supplementary Figures

**Supplementary Figure 1.**
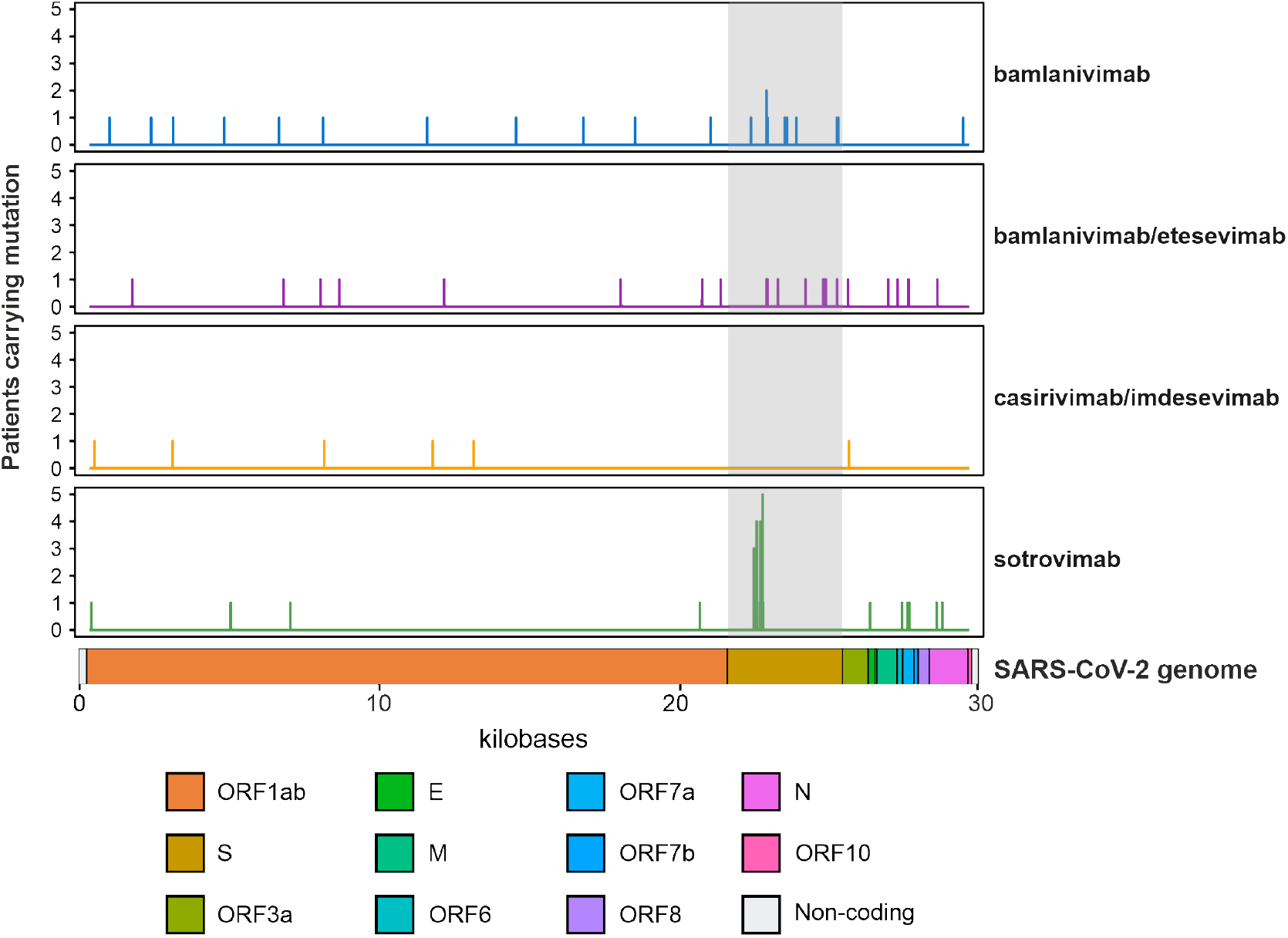
Patients receiving mAb treatment develop non-synonymous *de novo* mutations in the SARS-CoV-2 genome two (D2) to seven days (D7) after mAb infusion. Number of events of *de novo* mutations identified at D2 or D7 compared to D0 (baseline) are plotted across the positions in the SARS-CoV-2 genome. The number of patients developing mutations at specific positions in the SARS-CoV-2 genome are displayed for patients receiving bamlanivimab, bamlanivimab/etesevimab, casirivimab/imdevimab, and sotrovimab in the study. Only non-synonymous mutations are displayed in the figure. For more details, see **Supplementary Table 6**.

**Supplementary Figure 2.**
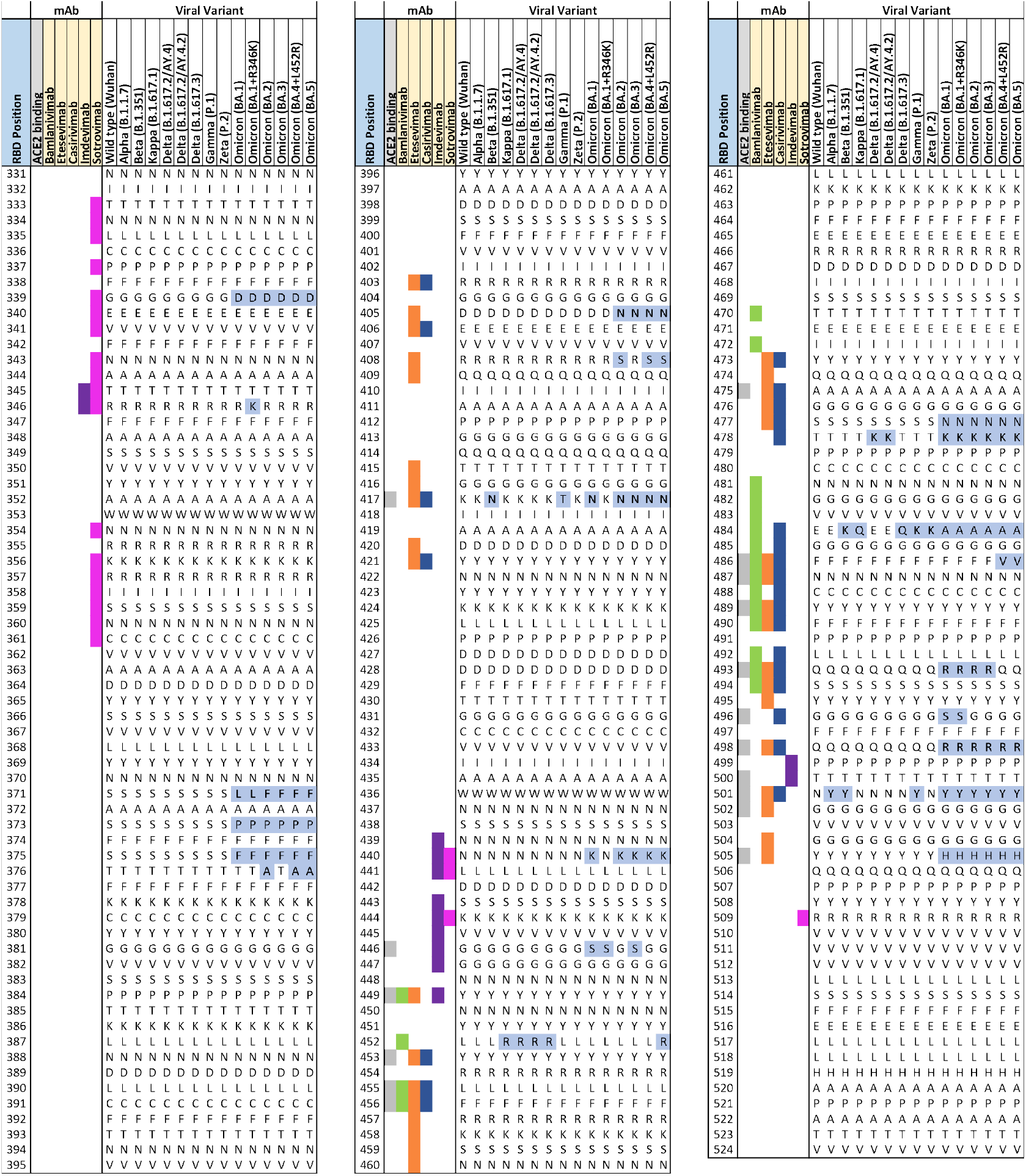
Multiple-sequence alignment of Spike RBD protein sequences of different SARS-CoV-2 variants and binding sites for human ACE2 (grey), bamlanivimab (green), etesevimab (orange), casirivimab (blue), imdevimab (purple), and sotrovimab (magenta). Spike RBD sequences from Wuhan (NC_045512), Alpha (B.1.1.7: EPI_ISL_674612), Beta (B.1.351: EPI_ISL_940877), Kappa (B.1.617.1: EPI_ISL_1384866), Delta (B.1.617.2/AY.4: EPI_ISL_1758376, B.1.617.2/AY.4.2: OX014422; B.1.617.3: MZ359842), Gamma (P.1: EPI_ISL_2777382), Zeta (P.2: EPI_ISL_717936), and Omicron (BA.1: EPI_ISL_6795848, BA.1.1: EPI_ISL_8724963, BA.2: EPI_ISL_8135710, BA.3: OM508650, BA.4+L452R: EPI_ISL_11542550, BA.5: EPI_ISL_11542604) are displayed. Non-synonymous amino acid residues compared to the Wuhan reference are highlighted in blue. Adapted from ref. (1).

**Supplementary Figure 3.**
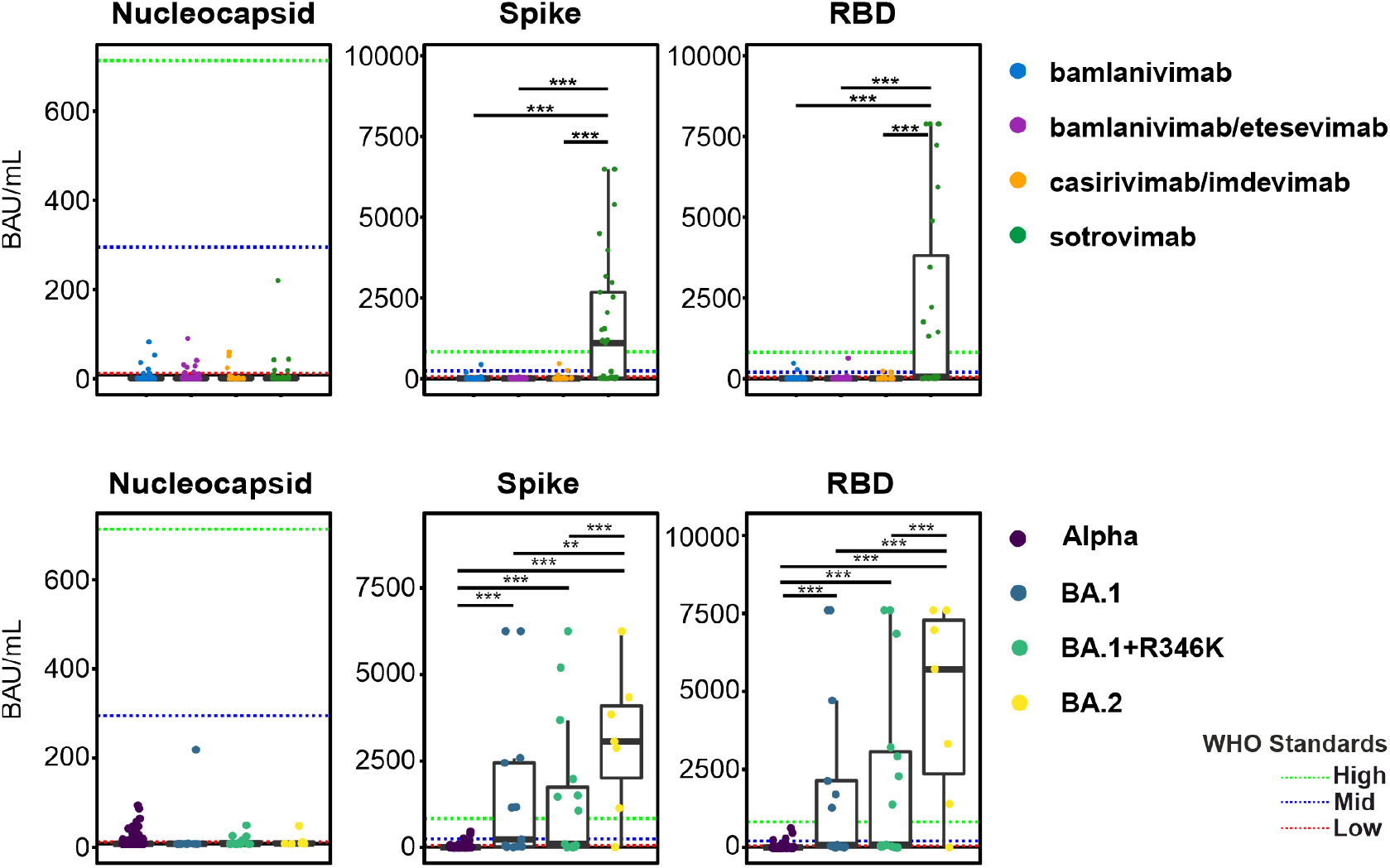
Anti-N, anti-S, and anti-RBD serology titers of patients receiving mAb therapy at D0 stratified by therapy and variant. Red, green, and blue dotted lines indicate SARS-CoV-2 WHO reference standard values for low, medium, and high antibody titers, respectively. *: *p* < 0.05. **: *p* < 0.01. ***: *p* < 0.001. D0: sample collected prior to mAb infusion.

**Supplementary Figure 4.**
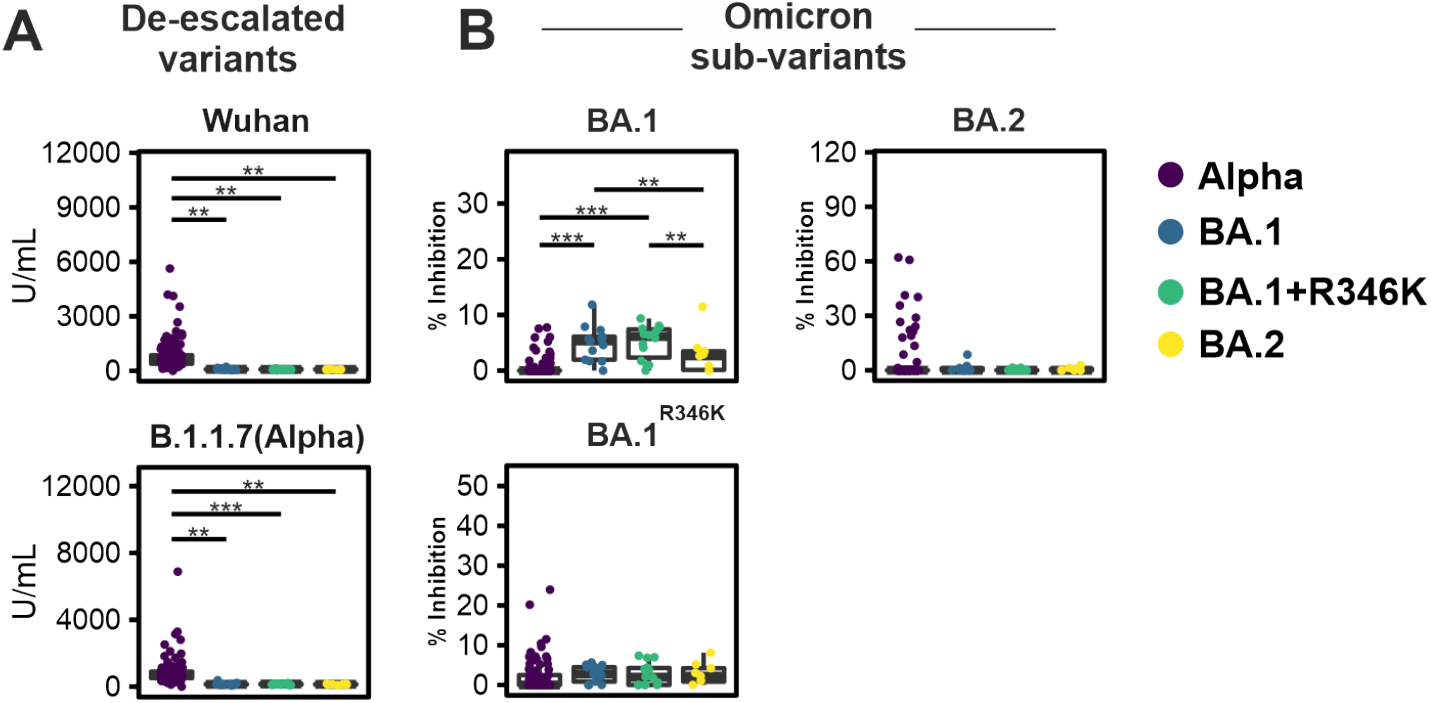
Anti-S neutralization capacity was measured against Alpha (B.1.1.7/Q4), BA.1, BA.1+R346K, and BA.2 variants at D2. Anti-S neutralizing antibody measurements against 5 different SARS-CoV-2 variants of concern in patients infected with Alpha, BA.1, BA.1+R346K, and BA.2 variants. *: *p* < 0.05. **: *p* < 0.01. ***: *p* < 0.001; D2: 2 ± 1 days after mAb infusion.

**Supplementary Figure 5.**
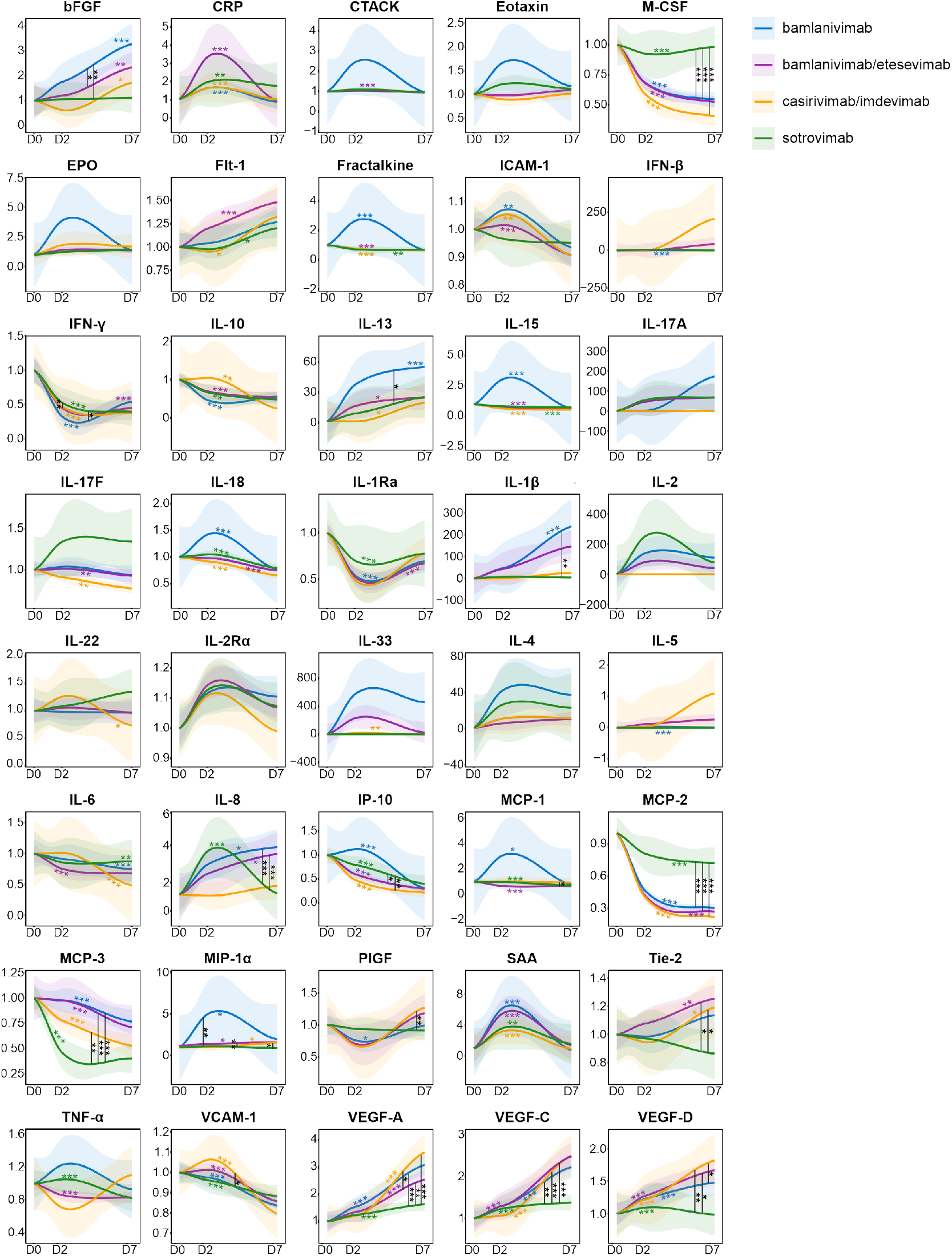
Temporal evolution of cytokines, chemokines, and growth factors (CCGs) in patients treated with bamlanivimab, bamlanivimab/etesevimab, casirivimab/imdevimab, or sotrovimab. Time is represented as days after mAb therapy (D0, D2, and D7). Colored askterisks in the graph refer to the significance of the slope from the 4 separate regression lines. Vertical lines with asterisks represent the significance of the pairwise comparison between the slopes in bamlanivimab, bamlanivimab/etesevimab, casirivimab/imdevimab, and sotrovimab therapy groups. *: *p* < 0.05, **: *p* < 0.01, ***: *p* < 0.001.

**Supplementary Figure 6.**
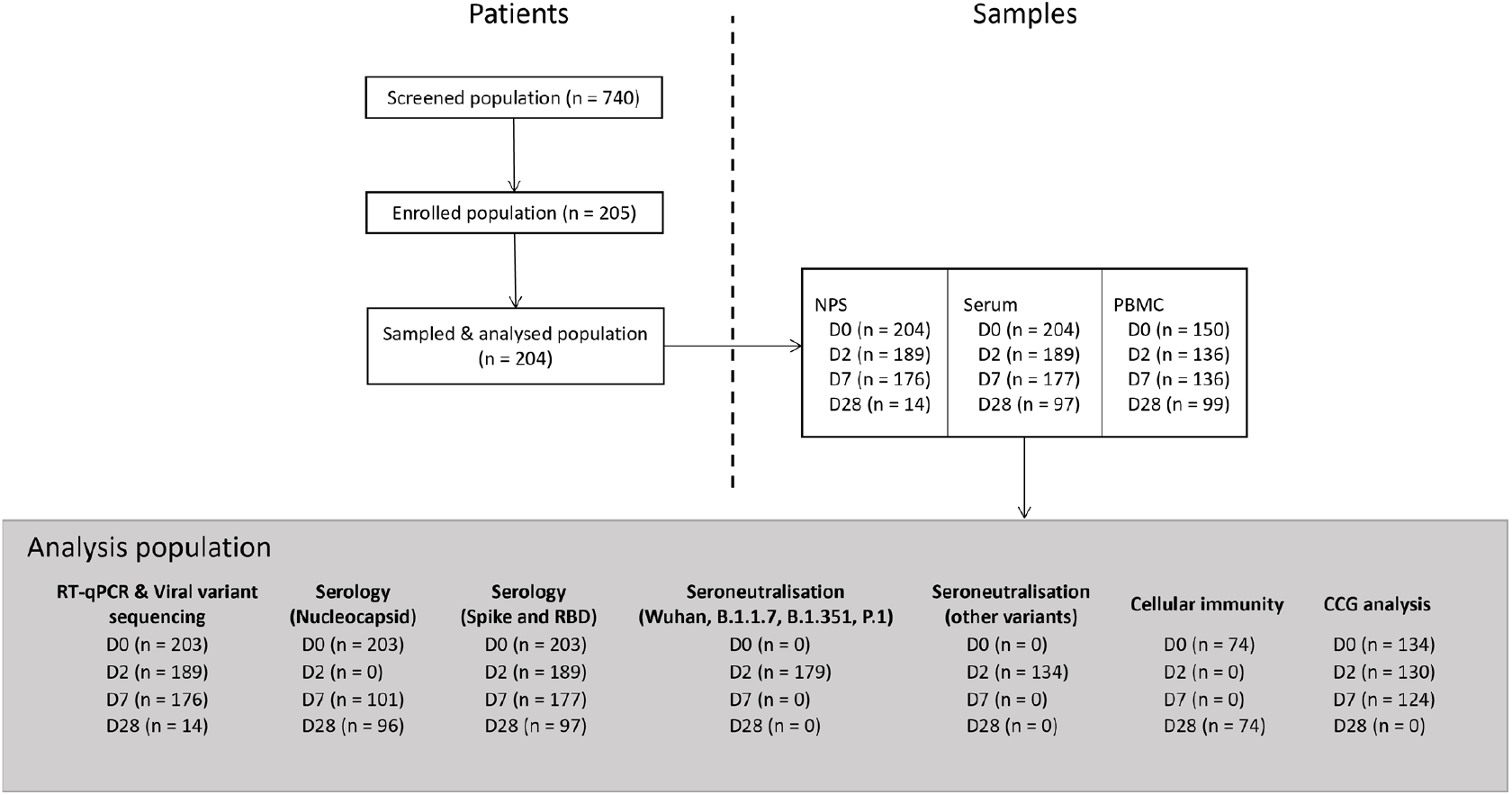
Overview of patient and sample inclusion in the study.

**Supplementary Figure 7.**
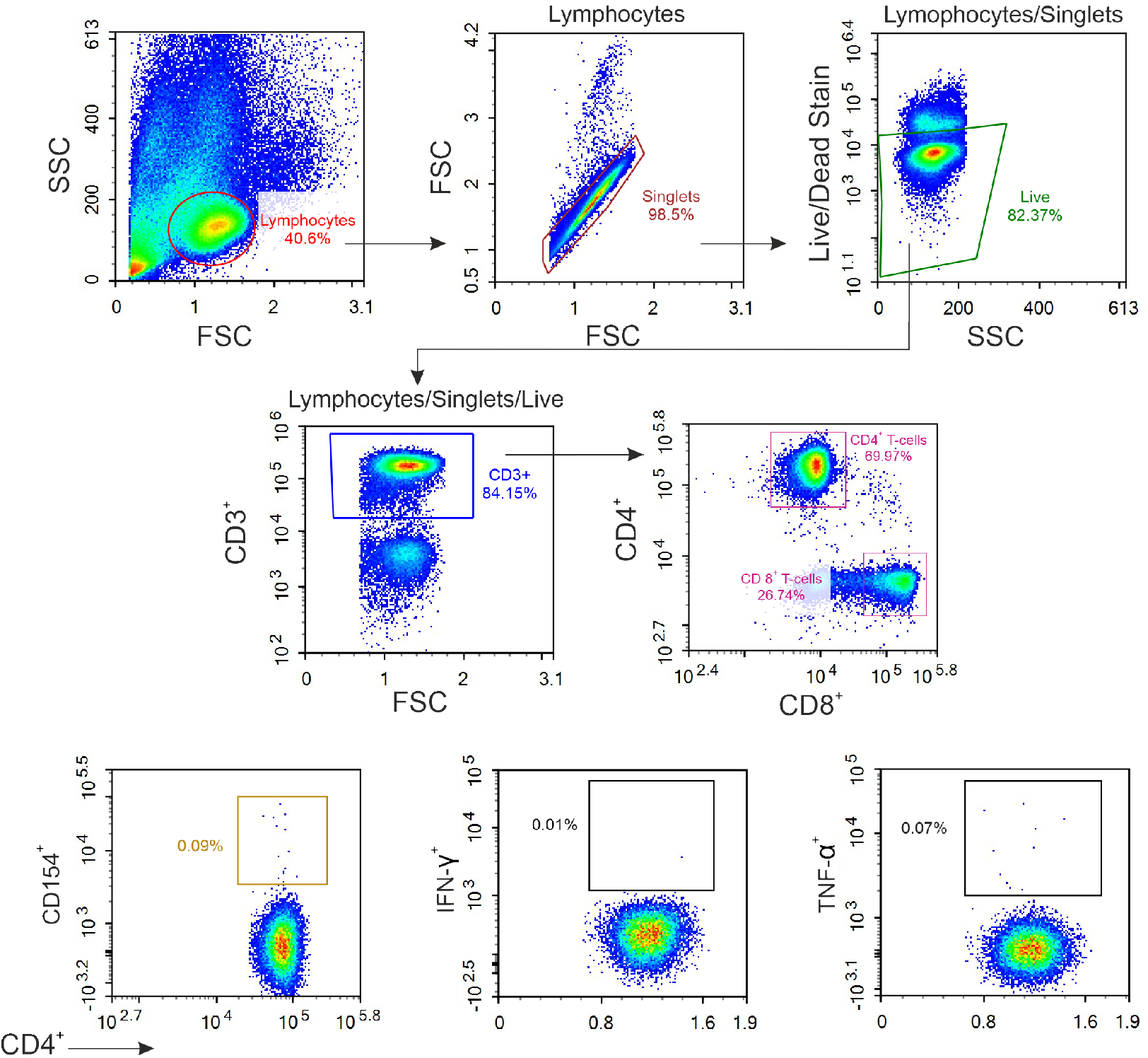
Representative flow cytometry plots for analysis of activated CD4^+^ T helper (Th) cells and their expression of effector cytokines. Gating strategy after specific stimulation with either a SARS-CoV-2 Nucleocapsid or a complete Spike peptide pool. PBMCs were gated on lymphocytes. Singlets were gated with dead cells excluded. Live CD3^+^ T cells were identified. Within the CD4^+^ Th cell populations activated CD154^+^ Th cells were gated, and the expression of IFN-γ and TNF-α analyzed. PBMC: Peripheral blood mononuclear cells.

### Supplementary Tables

**Supplementary Table 1:** Italian Medicines Agency Emergency Use Authorization eligibility criteria for bamlanivimab, bamlanivimab/etesevimab, casirivimab/imdevimab, and sotrovimab in adult patients as described by Savoldi *et al*.(2, 3).

|  |
| --- |
| **Patients enrolled during March 2021 – 15 June 2021:** |
| **All the following criteria should be met:**<br>1. Confirmed diagnosis of SARS-CoV-2 infection either by polymerase chain reaction or 3<sup>rd</sup> generation antigenic test on nasopharyngeal swab<br>2. Onset of at least one of the COVID-19 related symptoms among fever, cough, dyspnea, headache, myalgia, gastrointestinal symptoms, asthenia ≤ 10 days<br>3. Age ≥ 18 years<br>4. Body weight ≥ 40 kg<br>5. No need for oxygen therapy<br>6. No need for hospitalization<br>7. Presence of at least one of the following medical conditions:<br>• BMI ≥ 35 Kg/m<sup>2</sup><br>• Subject chronically undergoing peritoneal dialysis or hemodialysis<br>• Uncontrolled diabetes mellitus (HbA1c ≥ 9% or 75 mmol/L) or with chronic complications<br>• Primary immunodeficiency<br>• Secondary immunodeficiency (e.g., hematologic cancer patient in ongoing myeloid/immunosuppressive therapy or suspension for <6 months)<br>• Cardio-cerebrovascular disease (including arterial hypertension with documented organ damage) in subjects aged ≥ 55 years<br>• Chronic Obstructive Pulmonary Disease and/or other chronic respiratory disease in subjects ≥ 55 years |
| **Patients enrolled during 16 June – ongoing:** |
| **All the following criteria should be met:**<br>1. Confirmed diagnosis of SARS-CoV-2 infection either by polymerase chain reaction or 3<sup>rd</sup> generation antigenic test on nasopharyngeal swab<br>2. Onset of at least one of the COVID-19 related symptoms among fever, cough, dyspnea, headache, myalgia, gastrointestinal symptoms, asthenia ≤ 7 days<br>3. Age ≥ 18 years<br>4. Body weight ≥ 40 kg<br>5. No need for oxygen therapy<br>6. No need for hospitalization<br>7. Presence of at least one of the following medical conditions:<br>• Age > 65 years<br>• BMI ≥ 30 Kg/m<sup>2</sup><br>• Chronic kidney disease (including dialysis)<br>• Uncontrolled diabetes mellitus (HbA1c ≥ 9% or 75 mmol/L) or with chronic complications<br>• Primary immunodeficiency<br>• Secondary immunodeficiency (e.g., hematologic cancer patient in ongoing myelo/immunosuppressive therapy or suspension for <6 months)<br>• Cardio-cerebrovascular disease (including arterial hypertension with documented organ damage)<br>• Chronic Obstructive Pulmonary Disease and/or other chronic respiratory disease<br>• Chronic liver disease<br>• Hemoglobinopathies<br>• Neurodevelopmental diseases and neurodegenerative diseases |

**Supplementary Table 2.**
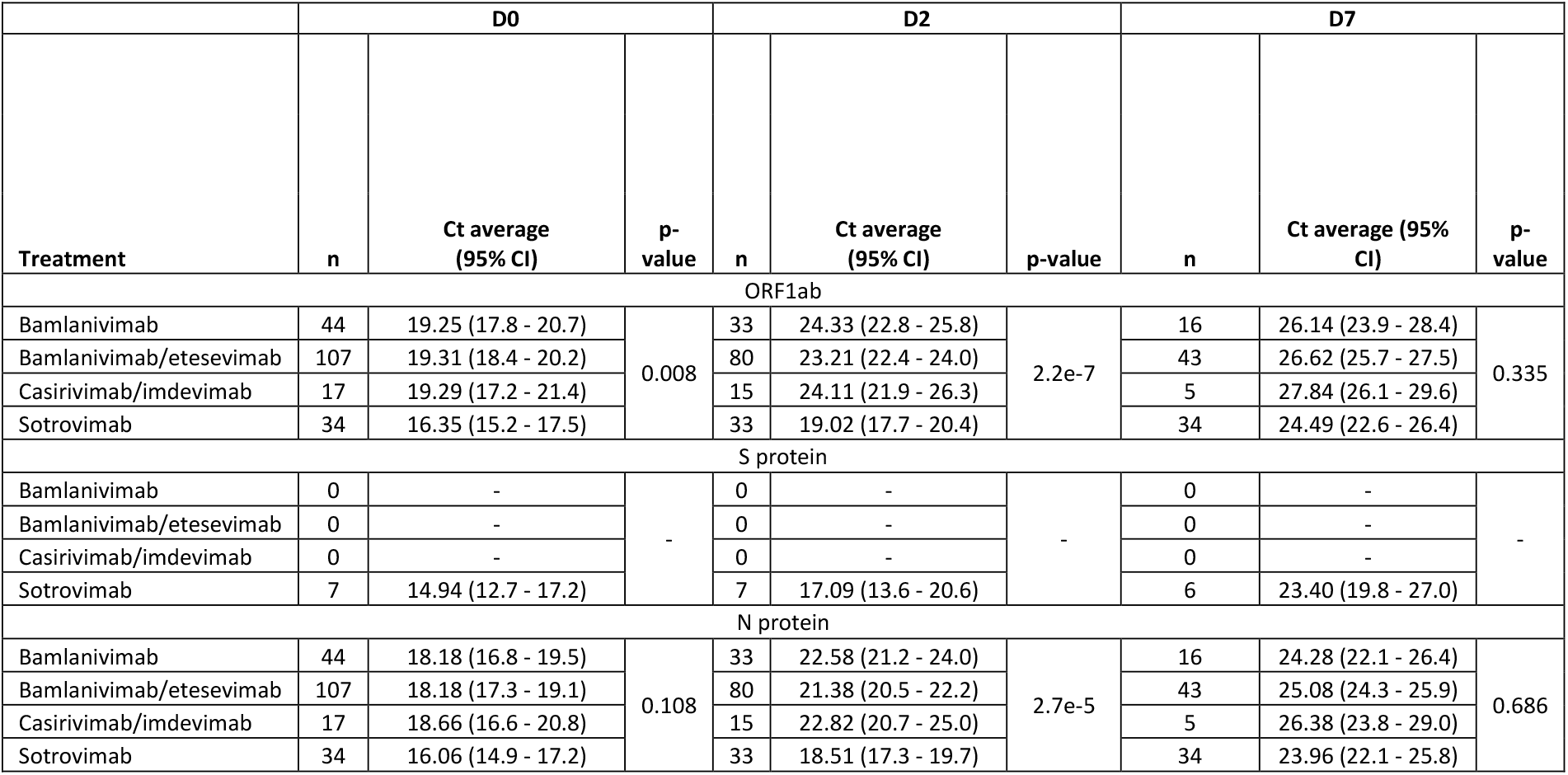
Results of real-time reverse transcriptase quantitative (RT-)qPCR detection of the ORF1ab, N, and S protein genes in nasopharyngeal swab samples collected from patients treated with bamlanivimab, bamlanivimab/etesevimab, casirivimab/imdevimab, or sotrovimab. Statistical comparisons between treatment groups at each timepoint were performed using Kruskal-Wallis. Total: total number of samples successfully analyzed by RT-qPCR. Ct: cyclic threshold. CI: confidence interval. (*): confidence intervals not calculated due to limited sample size. Only Ct values for variants of concern were considered. Positive samples collected at day 28 post mAb infusion were limited (2/14) and were therefore excluded from this analysis.

**Supplementary Table 3.** Results of real-time reverse transcriptase quantitative (RT-)qPCR detection of the ORF1ab, N, and S protein genes in nasopharyngeal swab samples collected from patients with different variants – Alpha and Omicron sub-variants. Statistical comparisons between treatment groups at each timepoint were performed using Kruskal-Wallis. Total: total number of samples successfully analyzed by RT-qPCR. Ct: cyclic threshold. CI: confidence interval. Positive samples collected at day 28 post mAb infusion were limited (2/14) and were therefore excluded from this analysis.

|  | D0 |  |  | D2 |  |  | D7 |  |  |
| --- | --- | --- | --- | --- | --- | --- | --- | --- | --- |
| Treatment | n | Ct average<br>(95% CI) | p-value | n | Ct average<br>(95% CI) | p-value | n | Ct average<br>(95% CI) | p-value |
| ORF1ab |  |  |  |  |  |  |  |  |  |
| Alpha | 161 | 19.3 (18.6 – 20.0) | 2.1e-4 | 123 | 23.5 (22.8 – 24.2) | 1.2e-8 | 62 | 26.5 (25.7 – 27.3) | 0.057 |
| Omicron | 34 | 15.1 (14.0 – 17.9) |  | 33 | 18.6 (16.4 – 21.4) |  | 34 | 23.6 (19.5 – 29.0) |  |
| B.1.1.7 | 146 | 19.4 (18.6 – 20.2) | 9.6e-4 | 110 | 23.5 (22.8 – 24.3) | 4.8e-7 | 56 | 26.9 (26.1 – 27.6) | 0.096 |
| Q.4 | 15 | 18.5 (16.2 – 20.9) |  | 13 | 23.4 (20.2 – 26.7) |  | 6 | 22.8 (17.8 – 27.9) |  |
| BA.1 | 13 | 18.2 (15.8 – 20.6) |  | 13 | 20.1 (17.3 – 22.9) |  | 13 | 25.0 (21.9 – 28.1) |  |
| BA.1+R346K | 14 | 15.3 (14.0 – 16.7) |  | 13 | 18.9 (17.2 – 20.6) |  | 14 | 23.9 (20.4 – 27.4) |  |
| BA.2 | 7 | 14.8 (14.1 – 15.1) |  | 7 | 16.1 (15.1 – 19.7) |  | 7 | 23.4 (21.7 – 27.3) |  |
| N gene |  |  |  |  |  |  |  |  |  |
| Alpha | 161 | 18.2 (17.5 – 18.9) | 0.008 | 123 | 21.7 (21.1 – 22.4) | 5.4e-6 | 62 | 24.9 (24.1 – 25.6) | 0.375 |
| Omicron | 34 | 15.3 (13.5 – 18.3) |  | 33 | 18.2 (16.0 – 20.7) |  | 34 | 23.0 (19.7 – 28.4) |  |
| B.1.1.7 | 146 | 18.3 (17.5 – 19.1) | 0.023 | 110 | 21.8 (21.1 – 22.5) | 1.2e-4 | 56 | 25.3 (24.6 – 25.9) | 0.248 |
| Q.4 | 15 | 17.4 (15.1 – 19.7) |  | 13 | 21.7 (18.7 – 24.6) |  | 6 | 21.2 (16.5 – 25.9) |  |
| BA.1 | 13 | 17.7 (15.3 – 20.2) |  | 13 | 19.3 (17.0 – 21.7) |  | 13 | 24.4 (21.3 – 27.5) |  |
| BA.1+R346K | 14 | 15.3 (13.8 – 16.8) |  | 13 | 18.6 (17.0 – 20.2) |  | 14 | 23.5 (20.0 – 27.0) |  |
| BA.2 | 7 | 14.4 (13.5 – 14.6) |  | 7 | 15.4 (14.4 – 19.4) |  | 7 | 23.0 (20.8 – 26.8) |  |

**Supplementary Table 4.** *De novo* SARS-CoV-2 variants emerging during the first seven days of monoclonal antibody treatment. Only non-synonymous mutations detected at D2 or D7 compared to D0 are reported. All reference positions refer to the Wuhan variant (GenBank: NC_045512.2). *: deletion. †: variant of concern mutation emerging irrespective of mAb therapy. Fs: frameshift.

| Reference position | ORF | Amino acid substitution | Previously reported mutations in the same codon |
| --- | --- | --- | --- |
| 701 | ORF1ab | G146S | — |
| 1478 | ORF1ab | A405S | — |
| 2841 | ORF1ab | V859A | — |
| 2864 | ORF1ab | E867* | — |
| 4592 | ORF1ab | E1443* | — |
| 6456 | ORF1ab | C2064Y | — |
| 6615 | ORF1ab | L2117S | — |
| 6843 | ORF1ab | S2193F | — |
| 7860 | ORF1ab | T2532I | — |
| 7987 | ORF1ab | Q2574H | — |
| 8505 | ORF1ab | T2747I | — |
| 11490 | ORF1ab | S3742F | — |
| 12067 | ORF1ab | M3934I | — |
| 13065 | ORF1ab | L4267S | — |
| 14503 | ORF1ab | H4747Y | — |
| 16795 | ORF1ab | V5511L | — |
| 18551 | ORF1ab | S6096T | — |
| 21458 | ORF1ab | I7065T | — |
| 22484 | S | V308L | — |
| 22578 | S | D339G | G339D(4) † |
| 22580 | S | E340K | E340K/A/D/G/Q(5-7) |
| 22581 | S | E340V | E340K/A/D/G/Q(5-7) |
| 22582 | S | E340D | E340K/A/D/G/Q(5-7) |
| 22599 | S | K346R | R346K/T/S/M(8) |
| 22673 | S | L371S | S371L, L371S(4, 9) † |
| 22679 | S | P373S | S373P, P373S(4, 9) † |
| 22686 | S | F375S | F375S(9) † |
| 22813 | S | N417K | K417N/T(4, 5, 10, 11) † |
| 22882 | S | K440N | N440K(8) |
| 23012 | S | E484K | E484K/Q(8, 12-15) |
| 23013 | S | E484A | — |
| 23014 | S | E484D | — |
| 23039 | S | Q493K | Q493K(8, 10, 11, 15) |
| 23040 | S | Q493R | Q493R(8, 10, 11, 15) |
| 23042 | S | S494P | S494P(12, 14) |
| 23401 | S | Q613H | — |
| 23709 | S | I716T | — |
| 24029 | S | F823I | — |
| 24939 | S | C1126F | — |
| 25024 | S | Y1155fs | — |
| 25407 | ORF3a | M5I | — |
| 25784 | ORF3a | W131L | — |
| 25811 | ORF3a | L140P | — |
| 26530 | M | D3G | — |
| 27145 | M | T208I | — |
| 27462 | ORF7a | C23W | — |
| 27610 | ORF7a | H73Y | — |
| 27874 | ORF7b | T40I | — |
| 28987 | N | Q239fs | — |

**Supplementary Table 5.** Distribution of patients tested for serological analysis among different treatment groups. Number and percentage of patients receiving bamlanivimab, bamlanivimab/etesevimab, casirivimab/imdevimab, or sotrovimab therapy that were fully vaccinated (14 days after second vaccination dose) or unvaccinated. Higher percentage of patients with high titers is observed in sotrovimab-treated patients, compared to other treatment groups.

| Therapy | Bamlanivimab |  |  | Bamlanivimab/etesevimab |  |  | Casirivimab/imdevimab |  |  | Sotrovimab |  |  |
| --- | --- | --- | --- | --- | --- | --- | --- | --- | --- | --- | --- | --- |
|  | n = 45 |  |  | n = 108 |  |  | n = 16 |  |  | n = 10 |  |  |
|  | Spike | RBD | Nucleocapsid | Spike | RBD | Nucleocapsid | Spike | RBD | Nucleocapsid | Spike | RBD | Nucleocapsid |
| Negative | 30 (66.6%) | 34 (75.5%) | 36 (80.0%) | 88 (81.5%) | 90 (83.3%) | 98 (90.7%) | 12 (75%) | 11 (68.8%) | 13 (81.3%) | 1 (10%) | 3 (30%) | 7 (70%) |
| Inconclusive | 9 (9.2%) | 7 (15.5%) | 3 (6.6%) | 18 (16.7%) | 15 (13.9%) | 2 (1.9%) | 2 (12.5%) | 2 (12.5%) | 0 (0%) | 6 (60%) | 3 (30%) | 0 (0%) |
| Positive – Low | 2 (4.4%) | 3 (6.6%) | 4 (8.8%) | 2 (1.9%) | 2 (1.9%) | 8 (7.4%) | 0 (0%) | 2 (12.5%) | 3 (18.6%) | 2 (20%) | 3 (30%) | 3 (30%) |
| Positive – Medium | 1 (4.4%) | 0 (0%) | 0 (0%) | 0 (0%) | 1 (0.9%) | 0 (0%) | 2 (12.5%) | 1 (6.2%) | 0 (0%) | 0 (0%) | 1 (10%) | 0 (0%) |
| Positive – High | 0 (0%) | 0 (0%) | 0 (0%) | 0 (0%) | 0 (0%) | 0 (0%) | 0 (0%) | 0 (0%) | 0 (0%) | 1 (10%) | 1 (10%) | 0 (0%) |
|  | n = 0 |  |  | n = 0 |  |  | n = 1 |  |  | n=25 |  |  |
|  | Spike | RBD | Nucleocapsid | Spike | RBD | Nucleocapsid | Spike | RBD | Nucleocapsid | Spike | RBD | Nucleocapsid |
| Negative | 0 (0%) | 0 (0%) | 0 (0%) | 0 (0%) | 0 (0%) | 0 (0%) | 0 (0%) | 0 (0%) | 1 (100%) | 3 (12%) | 2 (8%) | 22 (88%) |
| Inconclusive | 0 (0%) | 0 (0%) | 0 (0%) | 0 (0%) | 0 (0%) | 0 (0%) | 0 (0%) | 0 (0%) | 0 (0%) | 3 (12%) | 4 (16%) | 1 (4%) |
| Positive – Low | 0 (0%) | 0 (0%) | 0 (0%) | 0 (0%) | 0 (0%) | 0 (0%) | 1 (100%) | 1 (100%) | 0 (0%) | 1 (4%) | 1 (4%) | 2 (8%) |
| Positive – Medium | 0 (0%) | 0 (0%) | 0 (0%) | 0 (0%) | 0 (0%) | 0 (0%) | 0 (0%) | 0 (0%) | 0 (0%) | 0 (0%) | 0 (0%) | 0 (0%) |
| Positive – High | 0 (0%) | 0 (0%) | 0 (0%) | 0 (0%) | 0 (0%) | 0 (0%) | 0 (0%) | 0 (0%) | 0 (0%) | 18 (72%) | 18 (72%) | 0 (0%) |

**Supplementary Table 6.** SARS-CoV-2 Spike antigens of variants of concern (VOCs) tested by ACE2 seroneutralization in relation to the wild-type (Wuhan) SARS-CoV-2 variant. Amino acid modification and commonly used variant of concern (VOC) designations are summarized as described for the utilized V-PLEX® Serology Panel (Meso Scale Discovery (MSD), MD, USA) for VOCs and variants of interest used in this study.

| Lineages | Amino Acid Modifications | Common Designation |
| --- | --- | --- |
| B.1.1.7 | ΔH69-V70, ΔY144, N501Y, A570D, D614G, P681H, T716I, S982A, D1118H | Alpha |
| B.1.1.529; BA.1; BA.1.15 | A67V, ΔH69-V70, T95I, G142D, Δ143-145, Δ211/L212I, ins214EPE, G339D, S371L, S373P, S375F, K417N, N440K, G446S, S477N, T478K, E484A, Q493R, G496S, Q498R, N501Y, Y505H, T547K, D614G, H655Y, N679K, P681H, N764K, D796Y, N856K, Q954H, N969K, L981F | Omicron sub-lineage |
| B.1.1.529; BA.1+R346K; BA.1.1; BA.1.1.15 | A67V, Δ69-70, T95I, G142D/Δ143-145, Δ211/L212I, ins214EPE, G339D, R346K, S371L, S373P, S375F, K417N, N440K, G446S, S477N, T478K, E484A, Q493R, G496S, Q498R, N501Y, Y505H, T547K, D614G, H655Y, N679K, P681H, N764K, D796Y, N856K, Q954H, N969K, L981F | Omicron sub-lineage |
| B.1.1.529; BA.2; BA.2.1; BA.2.2; BA.2.3; BA.2.5; BA.2.6; BA.2.7; BA.2.8; BA.2.10; BA.2.12 | T19I, (L24-A27)toS, G142D, V213G, G339D, S371F, S373P, S375F, T376A, D405N, R408S, K417N, N440K, S477N, T478K, E484A, Q493R, Q498R, N501Y, Y505H, D614G, H655Y, N679K, P681H, N764K, D796Y, Q954H, N969K | Omicron sub-lineage |

